# Complex Genetics and Regulatory Drivers of Hypermobile Ehlers-Danlos Syndrome: Insights from Genome-Wide Association Study Meta-analysis

**DOI:** 10.1101/2025.09.19.25336146

**Authors:** Taylor Petrucci-Nelson, Sacha Guilhaumou, Takiy E Berrandou, Cortney Gensemer, Adrien Georges, Matthew Huff, Margaux-Alison Fustier, Asraa Esmael, Josephine Henry, Olivia Jaye, Ranan Phookan, Sarah Dooley, Kathryn Byerly, Brian Loizzi, Roman Fenner, Emma Mach, Amy Weintraub, Victoria Daylor, Julianna Weninger, Natalie Koren, Erika Bistran, Charlotte Griggs, Molly Griggs, Sydney Severance, Rebecca Byrd, Sunil Patel, Steven A Kautz, Anne Maitland, Nabila Bouatia-Naji, Russell A Norris

## Abstract

**Background:** Hypermobile Ehlers-Danlos syndrome (hEDS) is the most common subtype of EDS, a group of heritable connective tissue disorders. Clinically, hEDS is defined by generalized joint hypermobility and chronic musculoskeletal pain, but its impact extends beyond the musculoskeletal system. Affected individuals frequently experience autonomic, gastrointestinal, immune, and neuropsychiatric involvement, highlighting both the multisystemic nature of the condition and challenges of diagnosis. In contrast to other EDS subtypes with defined genetic causes, the molecular basis of hEDS has remained elusive.

**Methods:** We conducted a genome-wide association study (GWAS) of hEDS across three case controls studies, including 1,815 cases and 5,008 ancestry-matched controls. Fixed-effects meta-analysis of 6.2 million variants was complemented with LDAK gene-based association testing, transcriptome-wide association studies, and integrative annotation across multiple tissues and cell types including eQTLs, enhancer marks and open chromatin accessibility profiles, supported by luciferase assays on one candidate variant. LD-score genetic correlations were assessed between hEDS and 19 frequently reported comorbid conditions.

**Results:** Two loci reached genome-wide significance, including a regulatory region near the atypical chemokine receptor 3 gene (*ACKR3*) on chromosome 2. Functional annotation supports *ACKR3* risk alleles colocalize with eQTLs in tibial nerve, alter enhancer activity, and generate a *de novo* AHR transcription factor regulatory site, implicating neuroimmune and pain signaling pathways. Gene-based and transcriptome-wide analyses identified common variants in a locus containing multiple candidates, including *SLC39A13*, a zinc transporter critical for connective tissue development previously implicated in a rare form of EDS, and *PSMC3*, a gene involved in central nervous system development. LD-score regression revealed significant genetic correlations between hEDS and joint hypermobility, myalgic encephalomyelitis/chronic fatigue syndrome, fibromyalgia, depression, anxiety, autism spectrum disorder, migraine, and gastrointestinal diseases.

**Conclusions:** These results establish the first evidence of common variant contributions to hEDS, supporting a complex, multisystem model involving neuroimmune–stromal dysregulation. Our findings add novel indications to hEDS pathogenesis and provide solid foundations for future molecular definition and therapeutic discovery.

## Introduction

Ehlers–Danlos syndromes (EDS) are a group of heritable connective tissue disorders characterized by variable combinations of joint hypermobility, skin hyperextensibility, and tissue fragility. Most of the fourteen currently recognized subtypes are caused by defined pathogenic variants in structural extracellular matrix (ECM) proteins, enzymes essential for collagen biosynthesis, or other ECM-associated components^1^. Recent population-based estimates suggest that hEDS is the most common EDS subtype and related hypermobility spectrum disorders (HSD) may affect as many as 1 in 500 individuals,^2^ making them among the most prevalent heritable connective tissue disorders worldwide.

Despite its prevalence, hEDS lacks a molecular diagnostic test and remains defined by complex and non-specific diagnostic criteria. hEDS is characterized by generalized joint hypermobility and chronic musculoskeletal pain, but also involves multiple systems. Patients often present with autonomic dysfunction, gastrointestinal dysmotility, immune dysregulation, fatigue, migraine, sleep disturbances, neuropsychiatric symptoms.^3,4^ Structural complications such as mitral valve prolapse, aortic dilation, cerebrospinal fluid leaks, cranio-cervical instability, and hernias are also reported^5–7^. A global survey of over 3,000 patients reported an average of 24 co-occurring conditions per individual and diagnostic delays of up to two decades,^4^ highlighting both the disease economic burden^8^ and the shortcomings of current diagnostic frameworks.

The molecular underpinnings of hEDS remain largely undefined. Candidate gene studies and small-scale sequencing efforts have been limited in scope and statistical power, capturing only a fraction of the disorder’s genetic complexity^9^. Recent genetic investigations have identified low frequency variants in members of the kallikrein (KLK) serine protease family, particularly *KLK15* involved in a familial form of hEDS^10^. However, the lack of large-scale, high-powered studies has limited progress in defining the genetic architecture of hEDS, which is likely complex and shaped by multiple common risk variants contributing to its multisystem and prevalent clinical manifestations.

In this first genome-wide association study (GWAS) meta-analysis of hEDS, including more than 1,800 cases and 5,000 controls, we leveraged single variant, gene-based and transcriptome-wide analyses that demonstrate that hEDS has a complex genetic architecture. By integrating genetic associations, in *silico* and *in vitro* functional explorations from relevant tissues, we prioritized several genes likely regulated in nerve, skin, and fibroblasts. These findings point to neuroimmune signaling and neurodevelopment as key mechanisms underlying hEDS clinical heterogeneity. We also demonstrate shared genetic architecture with common comorbidities, including chronic fatigue, migraine, depression, anxiety, and gastrointestinal complications.

## Methods

### Patients and control populations

Two case control studies (MUSC1 and MUSC2) were assembled from hEDS patients through a national registry managed at the Norris lab, Medical University of South Carolina, USA. The MUSC1 cohort initially included 988 patients, and MUSC2 included 576 subsequently recruited patients. Across both cohorts, the mean age at enrollment was 40 years (range: 13-80 years). Self-reported clinical information captured a broad spectrum of symptoms and comorbidities documented through questionnaires, (Supplementary Tables 1, 2)^11^. For each study, cases were genetically matched to population-based controls using PCAmatchR package^12^. Controls (n = 2,305 for MUSC1; n = 1,343 for MUSC2) were matched from the UK Biobank, after exclusion of potentially confounding connective tissue and vascular disorders (Supplementary Methods, Supplementary Figure 1, Supplementary Table 3). A third case control study was derived from the AllofUs Research Program using electronic health records-based case identification^13^. After applying identical inclusion and exclusion criteria and the same case control matching strategy, we identified 524 hEDS patients and 1,386 matched controls (Supplementary Figure 1 - Supplementary Table 1). Details about genotyping or sequencing, imputation and major quality-control procedures are summarized in Supplementary Figure 1, Supplementary Table 4, 5, with additional details provided in the Supplementary Methods. Across all three studies, participants were predominantly of European ancestry (Supplementary Figure 1). All studies were approved by their relevant institutional ethics committees, and informed consent was obtained from all participants or their legal representatives. Further details are provided in Supplementary methods.

### Genome-wide association meta-analysis

For each study, association testing used additive logistic regression in PLINK v2.0 ^14^, adjusting for sex and the first ten principal components. Summary statistics were harmonized on hg19 and combined by fixed-effects inverse-variance meta-analysis in METAL^15^. We retained SNPs present in all three studies after removing heterogeneous signals (Cochran’s Q p < 0.01), yielding 6,181,533 high-quality variants in the final meta-analytic set (Supplementary Methods). Genome-wide significance was defined at P < 5×10^−8^.

### Gene-based association analysis

We performed gene-level association testing using the LDAK-GBAT, which implements a linear mixed model to estimate the contribution of each gene to the phenotype^16^. The model incorporates standardized SNP genotypes (matrix X) and phenotypic values (Y), partitioning variance into genetic (σ^2^_g_) and environmental (σ^2^_e_) components under normality assumptions^16^. For this analysis, we used a reference panel of 10,000 unrelated UK Biobank participants^17^, comprising 12,832,458 imputed SNPs aligned to the hg19 genome build. Gene boundaries were defined using RefSeq (NCBI37.3)^18^, and only SNPs located within annotated gene start and end positions were retained. We applied stringent filtering criteria to the summary statistics, retaining variants with minor allele frequency (MAF) ≥0.01, heterogeneity P-value >0.01, presence across all contributing case-control cohorts, and alignment with the reference panel. After filtering, 6,003,452 SNPs remained; of these, 2,392,275 were genic and mapped to 17,441 autosomal genes. As recommended by the method developers, we applied the “Human Default Model” in LDAK-GBAT^16^, weighting SNP heritability by MAF according to the function [MAF_j_(1−MAF_j_)]^0.75^. Empirical P-values were computed by permutation testing with 10 permutations per gene to approximate the null distribution. To account for multiple comparisons, genome-wide significance was set at P<2.87×10^−6^, corresponding to a Bonferroni correction over 17,441 tested genes. To mitigate redundancy due to correlation among gene-based signals, clumping was performed using the LDAK-GBAT procedure based on estimated genetic contributions: gene pairs on the same chromosome with squared correlation (r^2^) > 0.1 were identified, and only the most significantly associated gene in each group was retained.

### Data visualization tools and methods

Unless otherwise noted, all plots were generated in R (v4.4.2), using following packages: ggplot2 (v3.5.2), rtracklayer (v1.66.0), ggrepel (v0.9.6), data.table (v1.14.8), dplyr (v1.1.4), tidyr (v1.3.1), readr (v2.1.5), colorspace(v2.1-1). LocusZoom (http://locuszoom.org/) was used to provide regional visualization of GWAS results.

### Variant annotation

To identify the regulatory context of hEDS-associated variants, we used Integrated Genome Browser (IGB, v10.1.0) to visualize read density profiles and peak positions relative to the human reference genome^19^. Single nuclei ATAC-Seq datasets were retrieved from Human Enhancer Atlas (http://catlas.org/humanenhancer)^20^ and the Human Brain Enhancer Atlas (https://catlas.org/catlas_humanbrain/)^21^. Additional histone ChIP-Seq, bulk ATAC-seq and bulk DNAse-seq datasets were retrieved from ENCODE (https://www.encodeproject.org/)^22^. A complete list of datasets used in these analyses is provided in Supplementary Table 6. To predict transcription binding sites (TFBS), a 51 bp sequence centered on rs2600746 on chromosome 2 was used as input to the PERFECTOS-APE webserver (opera.autosome.org/perfectosape/scan), using HOCOMOCO (v13) core collection of position weight matrices as reference database^23^. The predicted structure of SLC39A13 was retrieved and visualized from AlphaFold Protein Structure Database (https://alphafold.ebi.ac.uk/).

### eQTL Lookup and colocalization

GTEx database (v10) was queried with lead SNPs and functionally relevant variants at hEDS-associated, generating violin plots from the web portal (www.gtexportal.org). To perform colocalization, we retrieved all SNP-gene eQTL associations from the GTEx v10 release (www.gtexportal.org/home/datasets). Colocalization between hEDS association signals and eQTLs was evaluated using coloc R package (v5.2.0), applying default prior values. Evidence for colocalization was considered when the posterior probability for shared association (H4) exceeded 80%. For each locus, the top candidate SNP was defined as the variant minimizing the product of eQTL and hEDS association *P*-values. Linkage disequilibrium (LD) with the lead SNPs was estimated in the European reference panel of the 1000 Genomes Project using the LDProxy function of the LDlinkR R package (v1.4.0).

### Transcriptome-wide association study (TWAS)

TWAS was performed using FUSION R/python package^24^. Gene expression reference models were pre-computed from the GTEx v8 dataset and provided by the original authors. Only genes with significant heritability P-value < 0.01 were retained for analysis. We evaluated genetically regulated expression across 15 Tissues from GTEx relevant to hEDS phenotypes: tibial nerve, transverse colon, sigmoid colon, skin (sun exposed lower leg), skin (not sun exposed suprapubic), thyroid, cells (cultured fibroblasts), tibial artery, coronary artery, aorta, whole blood, liver, lung, heart (left ventricle), heart (atrial_appendage). False discovery rate (FDR) correction was applied using p.adjust base function in R (v4.4.2).

### Mendelian Randomization (MR) of eQTLs

Significant SNP-gene eQTL associations and fine-mapped eQTLs from v10 release of GTEx database were retrieved from GTEx website (www.gtexportal.org/home/datasets) for the same 15 tissues mentioned above. To identify independent eQTLs matching with hEDS GWAS association study, for each gene and tissue significant, eQTLs were merged with hEDS GWAS. Top eQTL for each independent bloc (cs_id column) was then kept as instruments. Mendelian Randomization analysis was performed using TwoSampleMR R package (0.6.15), with eQTL as exposure and hEDS as outcome ^25^.

### Genetic correlation analysis

We estimated cross-trait genetic correlations (rg) between hEDS and a set of comorbid phenotypes frequently reported by patients, for which well-powered GWAS summary statistics were available. The selected traits included joint hypermobility, pain and autonomic diseases (myalgic encephalomyelitis/chronic fatigue syndrome, autonomic nervous system disorders, fibromyalgia and chronic pain), gastrointestinal diseases (irritable bowel syndrome, gastroparesis and gastroesophageal reflux disease), the following psychiatric or neurological diseases (migraine, major depressive disorder, autism spectrum disorder, and anxiety disorders), several hernias (abdominal, ventral, umbilical and inguinal hernias), commonly reported valve diseases (mitral valve prolapse and tricuspid valve disease), and pelvic organ prolapse (Supplementary Table 7). Summary statistics for each trait were harmonized to the hg19 reference genome, aligned to the same effect allele, filtered to HapMap3 SNPs, and reformatted with munge_sumstats.py function from LDSC software. Genetic correlations were then computed using LD Score Regression (LDSC v1.0.1) with European LD score reference files from the 1000 Genomes project. Analyses employed the standard bivariate model with an unconstrained intercept to account for sample overlap and population stratification. Standard errors were obtained by block *jackknife*. We pre-specified the multiple-testing correction as Bonferroni over 16 non-redundant phenotypes, grouping closely related outcomes (e.g., hernia subtypes) to avoid over-penalising highly overlapping constructs. The resulting significance threshold was α=0.05/16=3.1×10^−3^. For each trait, we report the genetic correlation coefficient (rg), its standard error, 95% confidence interval (CI) and nominal P-value. Further cohort-specific and QC details for each trait are provided in Supplementary Table 7.

### Luciferase reporter assays

A 3× tandem repeat of the genomic region surrounding the *ACKR3* lead SNP, rs2600746, encompassing either the protective or risk allele, was synthesized (VectorBuilder, Protective Allele: TCAGGCACATGCATCAGGCACATGCATCAGGCACATGCA, Risk Allele: TCACGCACATGCATCACGCACATGCATCACGCACATGCA) and cloned upstream of a minimal CMV promoter in a luciferase reporter vector. NIH3T3 cells were seeded at 10^5^ cells per well in 6-well tissue culture plates and transfected the following day with 2 µg total DNA per well using FuGENE® 4K transfection reagent (Promega, cat#E5911) at a 4:1 reagent:DNA ratio, according to the manufacturer’s instructions. At 72 h post-transfection, cells were lysed in 250 µl of Passive Lysis Buffer (PLB; Promega Dual-Luciferase® Reporter Assay System, cat#E1960). Lysates were cleared by centrifugation at 17,000 x g for 10 min at 4 °C, and 20 µl of supernatant was used per assay reaction. Firefly and Renilla luciferase activities were measured sequentially according to the manufacturer’s protocol on a luminescence plate reader. Each experimental condition was tested in *n* = 4 independent biological replicates, with 3 technical replicates per condition, and each well read in duplicate during the luciferase assay. Firefly activity was normalized to Renilla activity, and normalized ratios were expressed as fold change relative to the protective allele construct. Statistical significance was determined using one-way ANOVA with multiple pairwise comparisons.

## Results

### GWAS meta-analysis identifies two genetic risk loci for hEDS

We performed a GWAS meta-analyses across three case-control studies, comprising individuals predominantly of European ancestry (Supplementary Figure 1). After QC and ancestry matching, the combined dataset included 1815 cases and 5008 controls and involved 6,181,533 common variants (MAF>0.01) shared across all three studies.

Two loci achieved genome-wide significance for association with hEDS, located on chromosomes 2 and 8 (Figure 1A, Table 1, Supplementary Table 8). On chromosome 2, the lead variant rs2708184 was associated with an odds ratio (OR) of 1.29 (95% CI=1.18–1.41, P = 2.31×10^−8^, Table 1). Effect sizes estimates were concordant in all three case controls studies where the effect allele frequency (EAF) was 0.37 in cases, with no evidence for inter-study heterogeneity (P_Het_=0.83). On chromosome 8, the lead variant rs16880769 had an estimated odds ratio of 1.66 (95% CI= 1.42–1.93, P=1.33×10^−10^, EAF = 0.94), with consistent directions of effect and significance in all studies (P_Het_= 0.39, Table 1). Both GWAS signals were supported by clusters of correlated SNPs reaching genome-wide and suggestive levels of significance, providing robust evidence of association (Figure 1A, Supplementary Table 8).

**Table 1.**
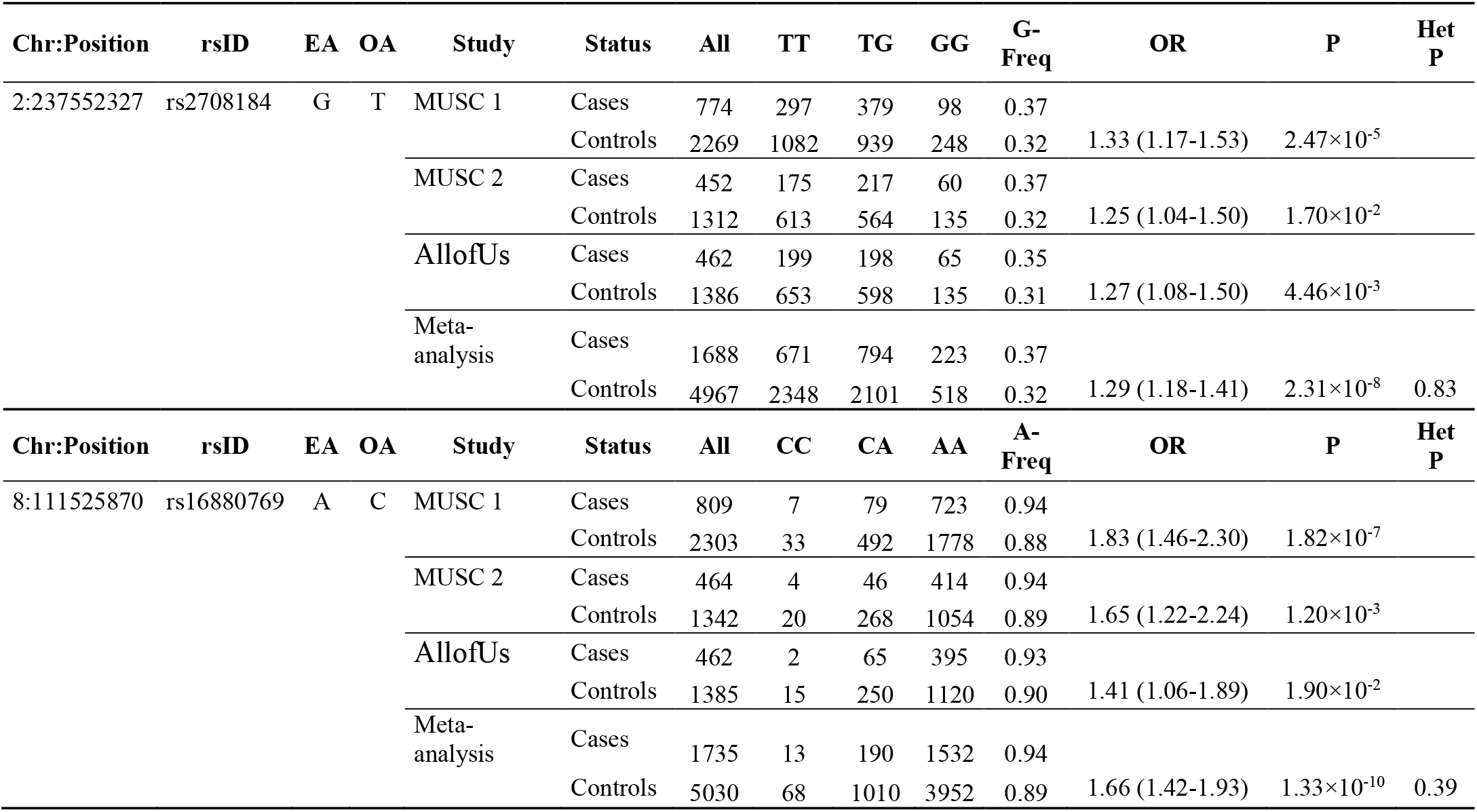
Lead associated variants at genome-wide significance in hEDS meta-analysis loci. Results are displayed for the lead SNPs of both genome-wide significant loci for the three GWAS case control studies (MUSC1, MUSC2, AllofUs) and in the GWAS meta-analysis. CHR: chromosome, POS: position (hg19), EA: effect allele, OA: other allele, N: total number of individuals, HOM_OA: homozygous for the other allele, HET: heterozygous, HOM_EA: homozygous for the effect allele, EAF: effect allele frequency, OR: odds ratio, P: Unadjusted P value of association obtained by two-sided Wald test, Het P: P values from the Cochran’s Q test for heterogeneity between studies in meta-analysis.

**Figure 1.**
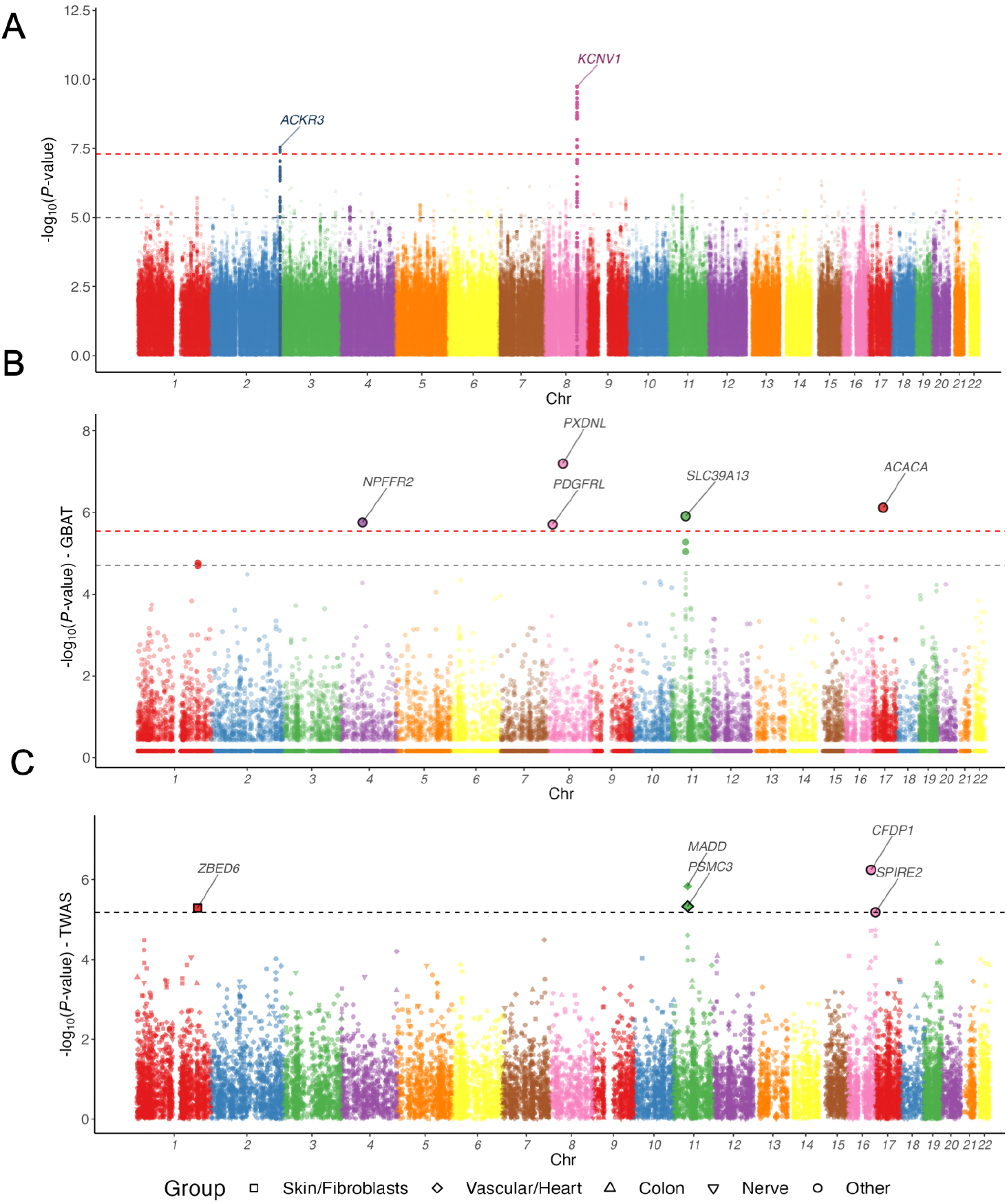
SNP and gene-based case control GWAS for hypermobile Elhers Danlos syndrome. **A:** Manhattan plot representation of meta-analysis of three case control GWAS including 1700 cases of hypermobile Elhers Danlos syndrome (hEDS) and 5000 controls. −log_10_ of association *P*-value (from a two-sided Wald test) is represented on the *y-*axis, genomic coordinates on the *x-*axis. SNPs located +/-500kb of genome-wide significant signals are highlighted. Closest gene of independent lead SNPs with *P*-value ≤ 5**×**10^−8^ is indicated for each locus. Dashed red line: *P*-value = 5×10^−8^. Dashed grey line: *P*-value = 1×10^−5^. **B:** Manhattan plot representation of gene-based case control GWAS using LDAK GBAT package. −log_10_ of association *P*-value (from a two-sided Wald test) is represented on the *y-*axis for each gene, genomic coordinates on the *x-*axis. Enlarged dot size indicates genes with FDR < 0.05. Names of Bonferroni significant genes are indicated. Dashed red line corresponds to Bonferroni significant threshold: *P*-value = 2.8×10^−6^. Dashed grey line: FDR < 0.05. **C**: Manhattan plot representation of hEDS transcriptome-wide association study (TWAS). −log_10_ of association *P*-value is represented on the *y-*axis for each gene, genomic coordinates on the *x-*axis. Enlarged dot size indicates genes with FDR < 0.05, which are indicated. Black lining indicates associations for which genetic colocalization was found (PP.H4.abf > 0.8). Only top tissue is shown for each gene. Dot shape represent tissue type. Dashed grey line: FDR < 0.05

### Functional annotation implicates regulatory elements on chromosome 2 and identifies *ACKR3* as a candidate target gene

The GWAS signal on chromosome 2 comprised several non-coding variants located ~70kb downstream of the atypical chemokine receptor 3 gene (*ACKR3*), also known as *CXCR7*. ACKR3 encodes a β-arrestin–biased receptor that functions as a scavenger for CXCL12, and modulates CXCR4-dependent chemokine signaling (Figure 2A). To evaluate regulatory potential, we queried the GTEx database for rs2708184, the lead variant at this locus and its high-LD proxies, to check for expression quantitative trait loci (eQTL) in human tissues. Four significant eQTLs were identified for *ACKR3* expression, most prominently in tibial nerve (Figure 2B), and additionally in cultured fibroblasts, subcutaneous adipose tissue and tibial artery (Supplementary Figure 2). The *ACKR3* eQTL in tibial nerve colocalized with hEDS GWAS association, with a posterior probability of 86%, supporting important likelihood of shared causal variant (Figure 2B, Supplementary Table 9, Supplementary Fig 2).

**Figure 2.**
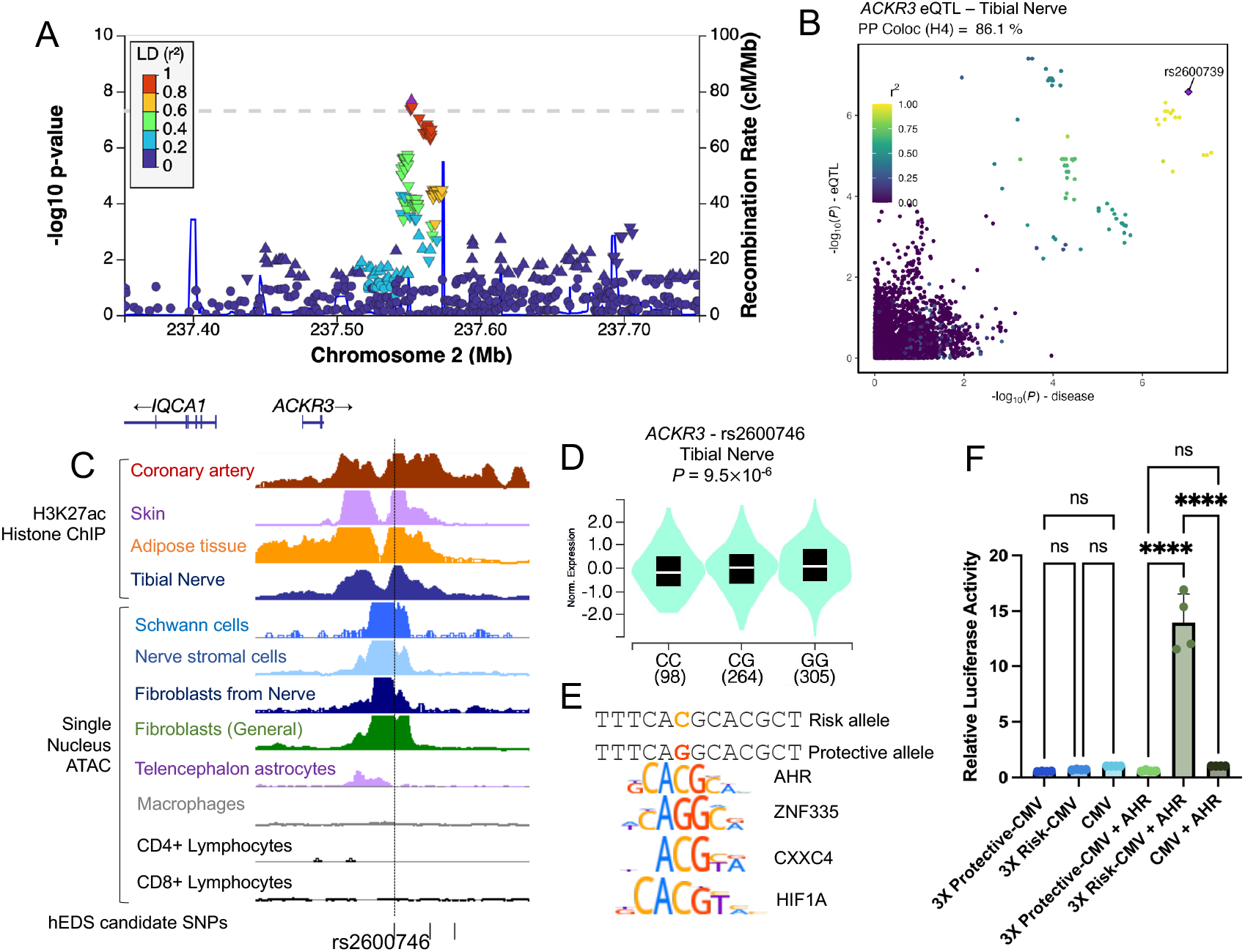
Gene prioritization and functional annotation at *ACKR3* locus. **A**. LocusZoom plot representing hEDS association at ACKR3 locus (chromosome 2). Dot shape indicate the lead SNP (diamond shape), and effect size (β) of nominally significant variants (upper triangle: β>0, lower triangle, β<-0, round shape: p-value>0.05). Dot-color indicates linkage desequilibrium (r2) with the lead SNP at each locus in the European subset of 1000G reference panel. **B**. x-y plot representing −log10(*P*-value) of *ACKR3* eQTL association in Tibial Nerve (*y*-axis) vs hEDS association (*x*-axis) for all SNPs within 500kb of lead SNP at *ACKR3* locus. Diamond dot shape indicates the SNP maximizing association to both *ACKR3* eQTL and hEDS. Dot-color indicates linkage desequilibrium (r2) with this SNP in the European subset of 1000G reference panel. Posterior probability for colocalization of both signals is indicated over the graph. **C**. Genome browser visualization of H3K27ac ChIP/ATAC-Seq/snATAC-Seq^20^ read densities in the region surrounding putative hEDS causal variants at *ACKR3* locus. Position of rs2600746 is indicated by dashed line. **D**. Violin plot representing the normalized expression of *ACKR3* in Tibial Nerve depending on rs2600746 genotype. Number of individuals for each genotype are indicated in legend. P-value of eQTL association is 9.5*×*10^−6^. **E**. Alignment of AHR, ZNF335, CXXC4 and HIF1A transcription factors motifs to rs2600746 C (risk) or G (protective) allele. Motif logos were retrieved from Hocomoco database (v13). **F**. A 3 times repeat of the genomic region containing either the protective or risk allele of rs2600746 was cloned upstream of a minimal CMV promoter and transfected into NIH3T3 cells. Firefly luciferase activity was normalized to Renilla luciferase and expressed as fold change relative to the protective allele construct. Data represent mean ± SD from n = 4 independent biological replicates with 3 technical replicates each. Statistical significance was determined by one-way ANOVA with multiple pairwise comparisons (α=0.05).

Inspection of epigenomic datasets from ENCODE and the Human Enhancer Atlas revealed strong enrichment for H3K27 acetylation, a histone modification characteristic of active regulatory regions in tibial nerve, among other tissue types near rs2600746, one of top hEDS associated variants in this locus (*P*=4.3 ×10^−8^, LD with lead SNP r^2^=0.97, Figure 2C). This genomic region also displayed high chromatin accessibility across multiple nerve-derived cell types, including fibroblasts, Schwann cells and stromal cells, as well as in telencephalon astrocytes, (Figure 2C). Together, these observations support a potential regulatory role for rs2600746 in tissues of direct relevance to hEDS clinical and biological manifestations. Consistent with this hypothesis, rs2600746 was a significant eQTL for *ACKR3* (*P*=9.5×10^−6^), with the hEDS risk allele (C) associated with reduced *ACKR3* expression in tibial nerve (Figure 2D). *In silico* transcription factor binding predictions indicated that rs2600746 alters several predicted binding sites, with the strongest signal was for the aryl hydrocarbon receptor (AHR), a widely expressed transcription factor; hEDS risk allele created a novel consensus binding site for AHR (Figure 2E, Supplementary Table 10). To experimentally validate these predictions, we performed luciferase reporter assays in fibroblasts that demonstrated that the rs2600746 risk allele conferred novel enhancer activity, and this effect was specifically dependent on the presence of AHR (Figure 2F).

Finally, although the chromosome 8 locus showed stronger statistical evidence for association, functional annotation of the credible set of variants revealed no clear regulatory features among the associated variants (e.g eQTLs associations, histone marks or chromatin accessibility) that would indicate regulatory activity or implicate a specific target gene. Nonetheless, based on its proximity to the signal (~540kb upstream of the lead variant) and its expression pattern in neural tissues, we suggest the potassium voltage-gated channel modifier subfamily V member 1 gene (*KCNV1*), as the most plausible candidate gene at this locus.

### Gene-based analysis supports the association of *SLC39A13* with hEDS

To increase discovery power beyond single-variant testing, we performed gene-based association analysis using GWAS summary statistics from the hEDS meta-analysis and the LDAK-GBAT framework.^16^ A total of 2,392,275 common variants mapping to 17,441 autosomal genes (RefSeq NCBI37.3) were retained. After clumping, five genes exceeded the Bonferroni-adjusted threshold for gene-based significance (P < 2.8×10^−6^, Figure 1B, Supplementary Table 11). Within these loci, the minimum single-variant p-values within each gene window ranged from 7.5×10^−7^ (*PDGFRL*) to 1.8×10^−4^ (*ACACA*), which contain suggestive but sub–genome-wide significant association signals, underscoring the gain in statistical power achieved by aggregating multiple modest SNPs effects into a gene-level test. Importantly, none of these genes were located within ±500 kb of the lead SNP-based GWAS signals on chromosomes 2 and 8, indicating that the gene-based framework revealed additional independent risk loci.

Among the top gene associations, *SLC39A13* (P=1.8×10^−6^) emerged as particularly relevant to hEDS susceptibility (Figure 3A). *SLC39A13* encodes a zinc transporter critical for connective tissue development, and rare homozygous mutations cause the recessive spondylocheiro-dysplastic form of EDS ^26–28^. Examination of individual variants within *SLC39A13* highlighted a common missense variant, rs61897432 (p.Thr20Ala, P_GWAS_ = 4.8×10^−6^) located in the gene’s first coding exon (Figure 3B). AlphaFold structural modeling suggests that Thr20 contributes to an alpha-helix domain, though confidence in this region is low. Given its relatively high frequency (MAF estimation is 0.15 in hEDS cases, 0.13 in controls), p.Thr20Ala is predicted to be benign (Figure 3C), suggesting potential contribution of additional potentially regulatory variants to the gene-based association in *SLC39A13*.

**Figure 3.**
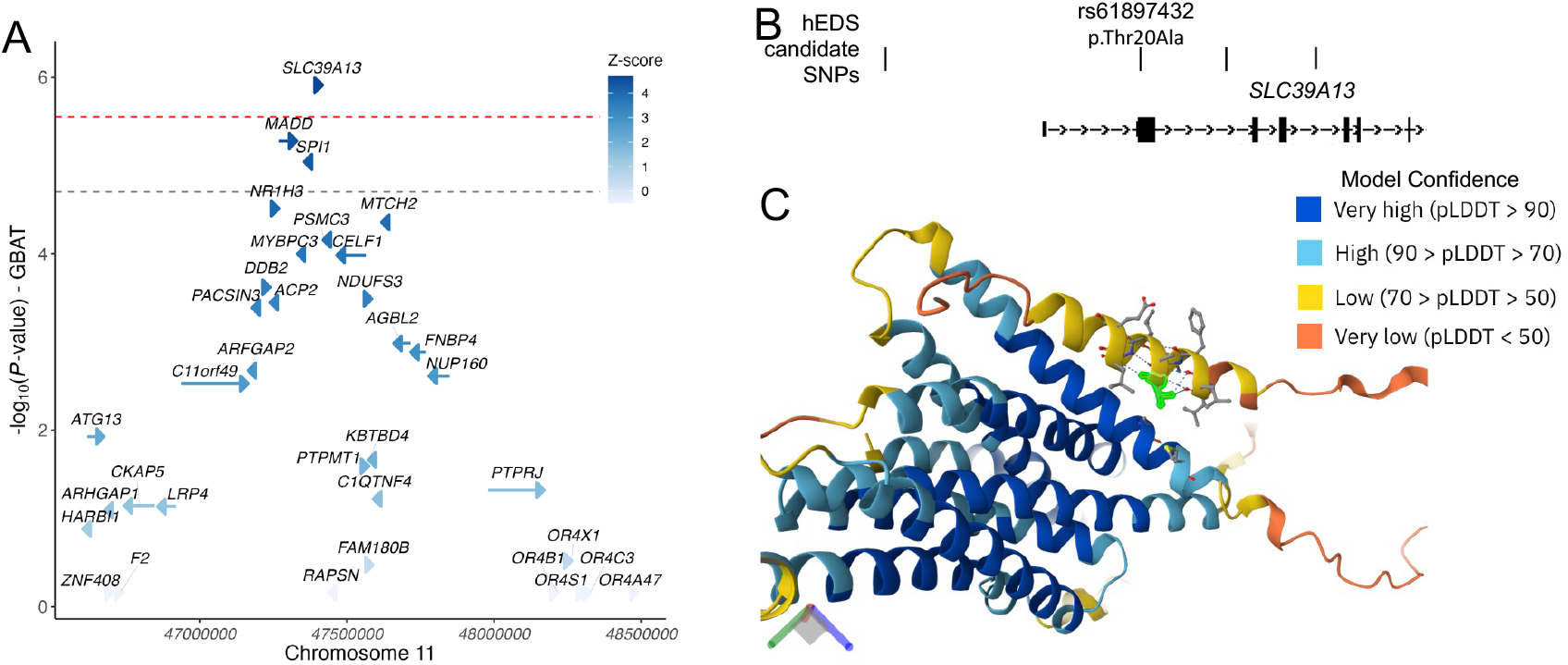
Gene prioritization and functional annotation at *SLC39A13* locus. **A**. Representation of hEDS gene-based association at *SLC39A13* locus (chromosome 11). −log_10_ of association *P*-value (from a two-sided Wald test) is represented on the *y-*axis. Arrows represent gene position and orientation along the genome (*x*-axis). Arrow color indicates Z-score of gene-based association. Dashed red line corresponds to Bonferroni significant threshold: *P*-value = 2.8×10^−6^. Dashed grey line: FDR < 0.05. **B**. Illustration of the position of candidate causal variants with respect to SLC39A13 transcription unit. Position of missense variant rs61897432 (p.Thr.20Ala.) is indicated. **C**. Representation of predicted protein structure for SLC39A13 and position of residue affected by missense variant. Structure prediction was retrieved and visualized from AlphaFold protein structure database (alphafold.ebi.ac.uk/). Backbone color indicates the confidence of structure prediction.

### Transcriptome-wide analyses identify genetically regulated expressed genes in relevant tissues to hEDS risk

To assess how genetic variation influencing gene expression contributes to hEDS susceptibility, we integrated GWAS summary statistics with eQTL data from GTEx using a transcriptome-wide association studies (TWAS) framework. Analyses focused on tissues relevant to hEDS biology, including skin, nerve, cultured fibroblasts, vasculature, colon, heart and blood (See Methods for full tissue list). Across these tissues, we identified five genes, whose genetically regulated expression was significantly associated with hEDS risk in at least one tissue (Figure 1C, Supplementary Table 12). The top signal was observed for the craniofacial development protein 1 gene (*CFDP1*), which showed consistent eQTL association across all analyzed tissues including blood, nerve, lung, heart, arteries, colon and skin (Figure 1C, Supplementary Table 12). Mendelian Randomization (MR) framework that uses top eQTLs as instruments to test for causal association, confirmed the correlation of higher expression of *CFDP1* with increased risk for hEDS, with strong evidence that the same allele drives both the GWAS and eQTL signals (posterior probability > 88% for colocalizing signals, Supplementary Table 12).

We also observed consistent TWAS and MR associations in the MAP kinase activating death domain gene (*MADD*), involving eQTLs in artery, colon and blood, with high posterior probably (87%) of a shared causal variant to increase *MADD* expression and hEDS risk only in blood (Supplementary Table 12). In addition, we found evidence from the TWAS and MR for the association of variants with decreased levels of expression of the proteasome 26S subunit, ATPase 3 gene (*PSMC3*) in heart and skin with higher risk for hEDS, and a high posterior probability (>94) to involve the same causal allele in both tissues.

Notably. *MADD* and *PSMC3* also demonstrated sub-genome-wide but suggestive significance in gene-based association testing (*P*=5.3×10^−6^ and *P*=7.0×10^−7^, respectively, Supplementary Table 11). Together with *SLC39A13*, these genes cluster in a genomic interval on chromosome 11 (Chr11:47.29–47.44 Mb). These association signals detected in the gene-based association and TWAS are driven by highly correlated variants (r2>0.90 in EU populations from HapMap). As such, this feature invalidated performing conditioned regression analyses to disentangle potential genetically independent effects of the three common variants.

### Shared genetics between hEDS and comorbid conditions

Patients with hEDS frequently report a broad range of comorbidities, spanning chronic pain and fatigue, neurological and psychiatric disorders, gastrointestinal dysfunction, and cardiovascular complications^11^. To assess the extent to which hEDS may present shared genetic and biological bases with some of these comorbidities, we retrieved and harmonized publicly available GWAS summary statistics for 19 diseases or traits (Supplementary Table 7) to conduct genetic correlation analyses with hEDS GWAS meta-analysis. As expected, hEDS shared a substantial proportion of its genetics with joint hypermobility (rg=0.41, P=8.0×10^−4^), its most reported clinical feature (Figure 4, Supplementary Tab 13). The most significant genetic correlation was found with myalgic encephalomyelitis/chronic fatigue syndrome (rg=0.35, P=6.5×10^−15^,), a condition reported by ~20 to 30% of patients included in our meta-analysis (Supplementary Table 2). We also observed a robust genetic correlation with chronic pain (rg=0.33, P = 5.16×10^−12^; reported by 73% to 91% patients), and to a less extent and significance with fibromyalgia (rg=0.21, P=0.043). Consistent with the high burden of gastrointestinal symptoms reported in >80 % patients, hEDS genetically correlated notably with irritable bowel syndrome (rg=0.31, P=1.71×10^−13^), gastroesophageal reflux disease (rg=0.18, P=2.87×10^−5^), and nominally with gastroparesis (rg=0.25, P=0.019).

**Figure 4:**
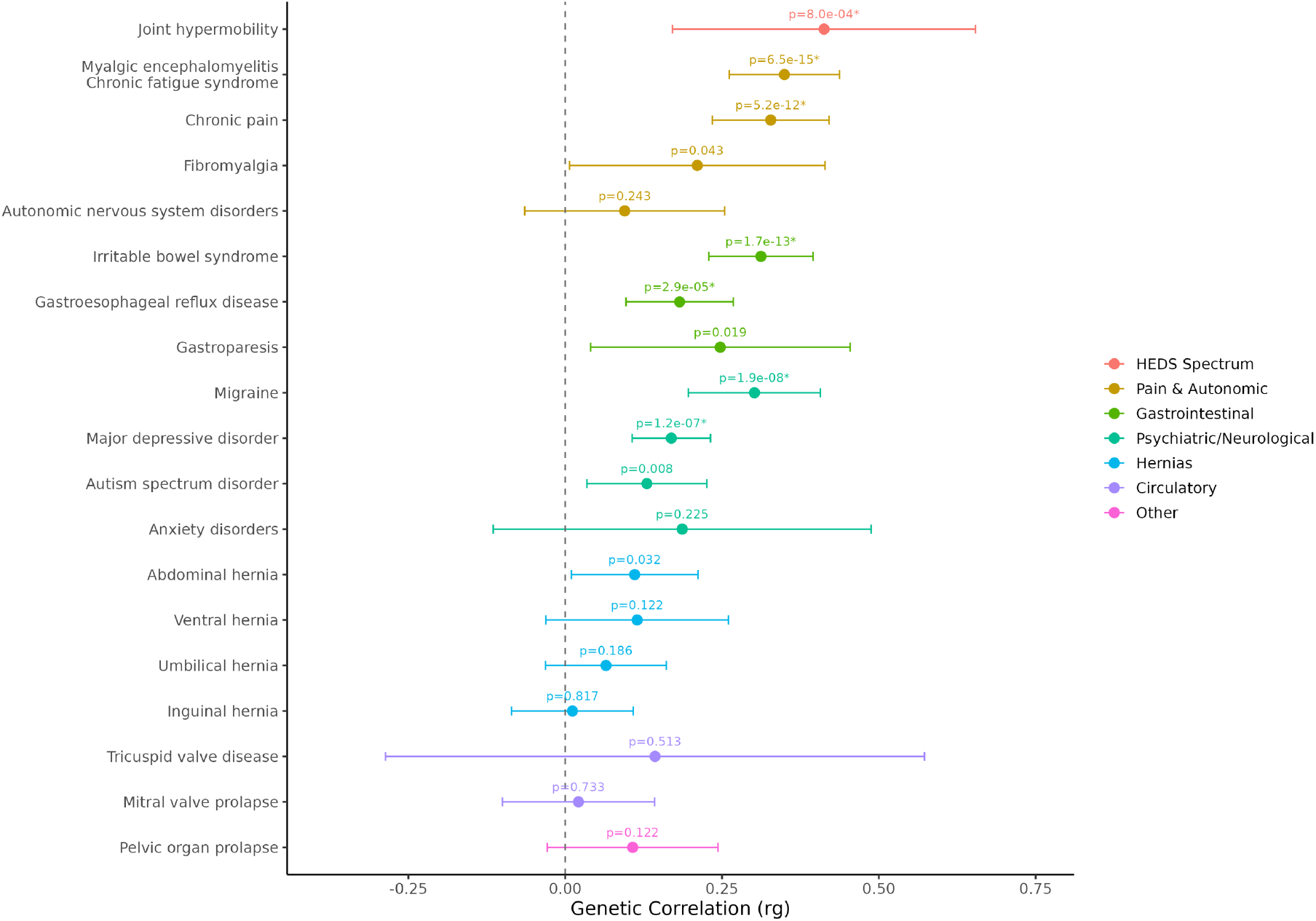
Genetic correlations between hEDS and frequently reported comorbidities in patients. Forest plot points show rg estimates with horizontal bars as their 95% confidence intervals. Traits are grouped by disease category and ordered by P-value within each category. The dashed line marks rg=0. Nominal P-values are displayed; associations meeting Bonferroni-corrected significance are marked with an asterisk and were defined as P<0.0031 considering 16 non-redundant phenotypes.

Beyond connective tissue and gastrointestinal track, hEDS showed significant shared genetic architecture with several neurological and psychiatric diseases including migraine (rg=0.30, P=1.9×10^−8^; reported by 50–68% of patients), major depressive disorder (rg=0.17, P=1.2×10^−7^; reported by 51 to 63% patients), nominally with autism spectrum disorder (rg=0.13, P=0.008, ~10% of patients) but not with anxiety (rg=0.19, P = 0.23), despite report by a large proportion of hEDS 64–76% of patients. Of note, our current analyses do not support large and significant genetic correlations between hEDS and structural or vascular manifestations including mitral valve prolapse, tricuspid valve disease, pelvic organ prolapse, or specific hernia subtypes (ventral, umbilical, inguinal), despite their frequent clinical reporting^11^ (Supplementary Table 2).

## Discussion

In this study, we provide a first genetic analysis of common variants contributing to hypermobile Ehlers-Danlos Syndrome (hEDS), the most common yet least genetically understood EDS subtype. Through a genome-wide association study (GWAS) meta-analysis leveraged for single variants, gene-based and transcriptome-wide associations, we demonstrate that hEDS present a complex genetic feature. By combining genetic association with functional annotations from diverse and disease-relevant tissues, we prioritized genes likely regulated in nerve, skin, and fibroblast populations. Biological insights from these loci converge on neuroimmune signaling and neurodevelopment, highlighting mechanisms that plausibly underlie the wide spectrum of clinical manifestations of hEDS. Furthermore, we demonstrate that hEDS shares substantial genetic architecture with joint hypermobility, its primary clinical feature, as well as with major comorbidities. These include gastrointestinal disorders such as irritable bowel syndrome, and neurological and psychiatric conditions, particularly migraine and major depressive disorder. Together, these findings provide the first molecular framework linking common genetic variation to hEDS susceptibility and substantially advance our understanding of its heterogeneous clinical presentation.

Deciphering the genetic basis of hEDS has proven particularly challenging, with prior studies yielding limited insights. Most genetic studies to date have implicated variants associated with other forms of EDS or connective tissue disorders, which rarely translate to hEDS-specific cohorts ^29,30^. Low frequency variants in the Kallikrein gene family were recently described in a couple of families, and mouse models carrying the hEDS risk *KLK15*^*G226D*^ allele recapitulate some core hEDS features, although the full multisystemic phenotype has yet to be fully evaluated^10^. Our findings extend this literature by demonstrating that, within a complex and suspected polygenic framework, common and primarily noncoding variants modestly increase hEDS risk, likely through regulatory effects on genes expressed in relevant tissues such as skin, nerve, and fibroblasts.

Among our most compelling findings is the identification of *ACKR3* within a validated genome-wide significant risk locus. ACKR3 encodes a β-arrestin–biased receptor that scavenges CXCL12 and modulates CXCR4 signaling with high expression in fibroblasts^9^, endothelial cells in the gut^31^ and vasculature^31^ and glial populations^32^. Prior work has demonstrated key roles for *ACKR3* in allergic responses^33^, angiogenesis^34^ and blood pressure regulation^35^, barrier function and epithelial repair through inflammation in the gastrointestinal system^36^, stress pathways and neurogenesis in anxiety and depression^37^, autoimmune diseases^38^, and neuroinflammation and nociceptive signaling in pain centers in the brain^39^. Our genetic and functional data converge on a likely causal SNP that lies within a strong enhancer active in nerve stromal and glial cells, colocalizing with eQTLs influencing expression across nerve, immune, and glial tissues. Notably, the SNP creates a consensus binding site for the AHR/ARNT complex that we demonstrated here to be functional *in vitro*. Given that AHR/ARNT integrate signals from dietary metabolites, xenobiotics, microbial products, and inflammatory cytokines^40^, the risk allele may confer context-dependent hEDS onset, consistent with patient-reported symptom flares that may be triggered by infections, trauma, or hormonal shifts. Beyond chemokine and response to external triggers, ACKR3 also scavenges endogenous opioid peptide such as nociceptin and dynorphin, thereby modulating their ability to stimulate classical opioid receptors^39^. Unlike canonical opioid receptors, ACKR3 does not engage G-protein signaling but regulates peptide bioavailability, positioning it as a potential modulator of pain thresholds, analgesic response, and opioid sensitivity, all frequently altered in hEDS. Taken together, these findings implicate ACKR3 variation as a biologically plausible driver of core hEDS disease mechanisms, particularly chronic pain, mast cell function in response to external or allergic stimuli, and neuroimmune dysregulation.

Through gene-based association analyses, we identified *SLC39A13*, a zinc transporter (ZIP13) previously implicated in the spondylodysplastic form of EDS, associated with hEDS for the first time^28^. Loss of function mutations in this gene causes a severe connective tissue phenotype, and *Zip13*-deficient mice display markedly reduced collagen content across multiple organs including the heart, kidney, intestine and skin^15^. ZIP13 is essential for connective tissue development and has been shown to gate an iron flux pathway that maintains iron homeostasis across several cellular compartments, a process directly relevant to collagen cross-linking during skin development.^41^ Our variant-level annotations highlighted p.Thr20Ala, a common coding variant within *SLC39A13*, which lacks clear predicted impact on protein function. Notably, this variant is in strong LD with several other SNPs that associate with hEDS through modulation of expression of two nearby genes: *MADD* and *PSMC3*. As a multifunctional adaptor protein that integrates TNF-receptor signaling with MAPK/ERK pathways, MADD is also a relevant candidate in hEDS given its established neuroprotective role.^42^ PSMC3, a 19S AAA-ATPase proteasome subunit, is critical for central nervous system development and has been linked to neurosensory syndrome in humans^43^ and rare neurodevelopmental disorders^44^. *PSMC3* pathogenic variants impair proteasome activity, triggering proteotoxic stress and dysregulation of immune and developmental signaling pathways, particularly through type I IFN signaling.^44^ Given the strong LD between coding and putative regulatory variants at this locus, statistical fine-mapping offered limited resolution. Future experimental studies will be necessary to identify the causal gene(s) and to elucidate the biological relevance of the multiple genetic signals detected.

This study provides evidence for shared genetic architecture between hEDS and several comorbid conditions frequently reported by patients, supporting potential shared genetic causes. Genetic correlation analyses revealed significant overlap with joint hypermobility, confirming that generalized ligamentous laxity and musculoskeletal features share common heritable determinants with hEDS. We also report large and significant genetic correlations with myalgic encephalomyelitis/chronic fatigue syndrome (ME/CFS), which affects up to 30% of hEDS patients, and chronic pain, and aligns with patient-reported fatigue as a central symptom^4^. Shared genetic contributions were also identified for gastrointestinal conditions such as irritable bowel syndrome and gastroesophageal reflux disease. Highly relevant findings also include the substantial genetic overlap between hEDS and several neuropsychiatric and pain-related disorders, including migraines, major depressive disorder and autism spectrum disorder. Notably, this overlap does not extend to anxiety, consistent with the heterogeneous clinical presentations observed in patients. Overall, our study provides genetic evidence of patient-reported multimorbidity and rationale for shared biological mechanisms underlying the multisystem burden of hEDS.

Our study has several important limitations. First, hEDS diagnosis remains complex and often requires years of multidisciplinary evaluation. Because most participants in our GWAS meta-analysis reported self-diagnosed hEDS, our findings may be influenced by diagnostic imprecision and enrichment for hypermobility spectrum disorders. Replication in clinically confirmed cohorts may refine these associations. Second, at the *SLC39A13* locus, strong LD among variants limited fine-mapping resolution, underscoring the need for functional validation and replication in larger, multi-ancestry datasets to improve signal localization for all reported hEDS GWAS loci. Third, our analyses of shared genetics with comorbidities were restricted to traits with sufficiently powered and currently available GWAS summary statistics, and future efforts should expand comparisons to a broader range of comorbidities to provide a more complete picture of shared genetic mechanisms.

In conclusion, while generalized joint hypermobility and tissue fragility remain hallmark features of hEDS, our genetic findings suggest a broader pathophysiological model, in which hEDS present features of a neuroimmune–stromal disorder, in which inherited variants disrupt regulatory pathways across connective tissue, neurodevelopmental pathways, immune, neurological, pain, and gastrointestinal systems. Regulatory variants at loci such as *ACKR3* and *SLC39A13*-MADD-*PSMC3* exemplify how context-dependent gene expression may contribute to symptom variability and disease severity. Fully defining the pathophysiology of hEDS, and ultimately advancing precision medicine, will require even larger and more deeply phenotyped cohorts, combined with integrative approaches that unify genomic, transcriptomic, behavioral and environmental triggers data. Such efforts are more likely to completely capture the dynamic, multisystem nature of this complex and understudied disorder.

## Data Availability

All data produced in the present study are available upon reasonable request to the authors.

## Acknowledgements

This work was made possible through the generous support of the Maltz Family Foundation, The Fullerton Foundation, The Connective Tissue Coalition, and the many individual donors who continue to stand behind our mission. Their contributions have been instrumental in advancing the understanding of hEDS and HSD, driving comprehensive survey-based research, enabling genetic and biomarker discoveries, and fueling the pursuit of innovative therapies. Most importantly, this support helps us move closer to improving outcomes for a patient community that is both commonly affected and too often underserved. This work was supported by the National Institutes of Health (NIH; grant number F32AI181339 to C.G.). T-E.B. is supported by a fellowship from the *Fondation pour La Recherche Medicale* (FRM). The Genotype-Tissue Expression (GTEx) Project was supported by the Common Fund of the Office of the Director of the National Institutes of Health, and by NCI, NHGRI, NHLBI, NIDA, NIMH, and NINDS. Genotyping was partially supported by the Spanish National Cancer Research Centre, in the Human Genotyping lab, a member of CeGen Biomolecular resources platform (PRB3), to be supported by grant PT17 /0019, of the PE I+D+i 2013-2016, funded by Instituto de Salud Carlos III and a European regional development fund (ERDF).

We acknowledge participants of the All of Us Research Program for their contributions, without whom this paper’s findings would be incomplete. We also thank the National Institute of Health for making the All of Us Research Program’s participant data available for examination in this study. This research has been conducted using the UK Biobank Resource under application number [91558].

## Author contributions

Writing or editing the manuscript: T. P.-N., S. G., T.-E. B., C. G., A. G., S. P., A.E., A. M., N. B.- N., R. A. N. Study design and conception: T. P.-N., T.-E. B., C. G., A.G., S. P., S. A. K., A. M., N. B.-N., R. A. N. Data collection: T. P.-N., C. G., R. F., A. W., V. D., J. W., N. K., E. B., C. G, M. G., S. S., R. B., R. A. N. Performed experiments: T. P.-N., C. G., M. H., O. J., R. P., S. D., K. B., B. L., E. M., N. B.-N. Data management: T. P-N., C. G., M. H., A. E. Statistical analysis: S. G., T.-E. B, M. H., A. G., M.-A. F., J. H. Study direction: N.B-N and R.A.N.

