## Supplementary methods, figures and tables for "Complex Genetics and Regulatory Drivers of Hypermobile Ehlers-Danlos Syndrome: Insights from Genome-Wide Association Study Meta-analysis"

##### MUSC /UKBB case control studies

From January 2021 to January 2023 (MUSC1) and February 2023-Jan 2024 (MUSC2), participants in this study were enrolled by clinical coordinators under Medical University of South Carolina (MUSC) Institutional Review Board (IRB) approval [Pro00098399]. During the consent process, participants provided written, informed consent, and responded to a series of clinical and demographic questions verbally. Participants were over the age of 12 and self-reported a clinical diagnosis of hEDS. Both males and females were included and gender, race and ethnicity were self-reported. Saliva samples were collected from enrolled participants using Oragene DNA saliva collection kits (DNA Genotek), which were mailed directly to participants after they enrolled in the study. Upon receipt, genomic DNA was extracted from 0.5 mL of saliva using the prepIT•L2P kit (DNA Genotek, Ottawa, Canada) according to the manufacturer's instructions with minor modifications. Briefly, saliva samples were incubated at 50 °C prior to extraction, treated with PT-L2P reagent, and centrifuged to remove debris. The supernatant was precipitated with ethanol, and DNA pellets were washed with 70% ethanol, air-dried, and resuspended in TE buffer. DNA concentration and purity were assessed using a NanoDrop spectrophotometer (Thermo Fisher Scientific), and samples were stored at −20 °C until use as two separate batches of patients samples (MUSC 1 and MUSC 2 cohorts).

MUSC 1 and MUSC 2 cases were genotyped on the Illumina Global Screening Array +MD v3.0 and imputed to the Haplotype Reference Consortium version 1.1 reference panel [1] on the Michigan Imputation Server [2]. Genotypes of the UK Biobank controls were drawn from the Axiom array resource with centrally provided imputation.

We leveraged participants from the UK Biobank resource to identify ethnically matched controls to MUSC patients. This research has been conducted using the UK Biobank Resource under application number [91558]. Participants provided informed, broad consent for their anonymized data and samples to be used in health-related research, in accordance with UK Biobank's Ethics and Governance Framework.

#### AllofUs case control study

We included genetic data from participants through the United States National Institute of Health (NIH) All of Us research program[3]. Access to the All of Us Controlled Tier dataset (version 7), available to authorized users through the Research Workbench, was granted through a Data Use and Registration Agreement (DURA) registered between MUSC and the NIH (https://www.researchallofus.org/institutional-agreements/). Participants were provided informed consent following a protocol reviewed by the All of Us Institutional Review Board (IRB), in accordance with the regulations and guidance of the NIH Office for Human Research Protections. Participants who consented to participate donated fresh whole blood as a primary source of DNA for sequencing. The AllofUs cohort used whole-genome sequencing (WGS; ACAF dataset). All analyses were aligned to hg19; AllofUs variants were converted from hg38→hg19 using liftOver v1.0.4 [4].

#### Case control selection and matching

To minimize confounding by ancestry, controls were matched to cases using the PCAmatchR package [5], which leverages Mahalanobis distance on principal components derived from ancestry-informative SNPs [5]. From UK Biobank we matched controls to MUSC 1 and to MUSC 2 (after exclusions for missingness/heterozygosity and ICD-based confounding diagnoses: Marfan Syndrome, Loez-Dietz, all forms of EDS, and other related diseases, Supplementary Table 3). We applied identical strategy to select controls from AllofUs to match the cases identified in this resource.

#### Per-study GWAS cohorts, QC and association models

##### Overview of cohorts

Step-wise sample exclusions and final per-study sample sizes are reported in Supplementary Table, Supplementary Figure 1). We removed individuals with missingness/heterozygosity (mean ± 3 SD), PCA-defined non-European ancestry (>6 SD from the 1000G European centroid), and related individuals (KING kinship > 0.0625). For UK Biobank controls, individuals with confounding diagnoses were additionally excluded by ICD codes. Variant-level milestones are provided in Supplementary Table 5). Post-imputation/WGS filters were harmonized across studies: autosomal biallelic SNPs with Rsq > 0.8 (for imputed datasets), MAF ≥ 0.01, missingness < 5%, and Hardy–Weinberg equilibrium $p \geq 1\times10^{-6}$ (evaluated in controls). The final per-study variant sets comprised 6,685,653 (MUSC 1), 6,609,472 (MUSC 2), and 7,918,868 (AllofUs) SNPs (Supplementary Table 5). After application of individuals and variants quality control criteria, 866 cases in MUSC1, 497 cases in MUSC2 and 462 cases in AllofUs were retained for analysis.

##### Association model

Each study was analyzed separately using additive logistic regression in PLINK v2.0 [6], adjusting for sex and the top ten ancestry principal components. Only high-quality autosomal biallelic SNPs were retained for the final per-study analyses (thresholds below).

***MUSC 1 case control study***. Among 988 enrolled cases, we removed 107 as PCA-defined non-European outliers (> 6 SD from the 1000G European centroid), and 12 for relatedness (KING kinship > 0.0625), leaving 869 cases prior to post-imputation checks. Three additional sample failed missingness/heterozygosity (mean ± 3 SD), yielding 866 cases. Controls were selected from a pool of ~100,000 UK Biobank Europeans using PCAmatchR [5] to form an ancestry-matched set of 2,601; 9 were removed for missingness/heterozygosity and 287 for ICD-based confounding diagnoses, leaving 2,305 controls.

Case genotypes started with 730,059 variants; keeping SNVs and non-monomorphic sites gave 599,610, duplicates removal 595,621, and a pre-imputation call-rate filter (< 2 % missing) 576,098; after HRC alignment 551,645 variants entered imputation. The imputed source comprised 40,355,712 variants; restricting to autosomal biallelic SNPs with Rsq > 0.8 and MAF ≥ 0.01 and intersecting across cohorts produced 7,021,115 variants. Post-imputation missingness (< 5 %) retained 6,686,384, and HWE in controls ($p \geq 1\times10^{-6}$) yielded 6,685,653 variants for analysis. The matched UK Biobank controls mirror these post-imputation counts.

***MUSC 2 case control study.*** Of 563 cases, 56 as PCA outliers and 4 for relatedness (kinship > 0.0625), leaving 503 prior to post-imputation checks; 6 further samples were removed for missingness/heterozygosity, yielding 497 cases. From a 100,000-person UK Biobank European pool, 1,506 controls were matched; 19 were excluded for missingness/heterozygosity and 144 for confounding diagnoses, leaving 1,343 controls. Variant processing began with 730,059 genotyped variants; SNV/non-monomorphic filtering left 587,181, duplicates removal 583,354, and a call-rate filter 553,424; 531,821 HRC-aligned variants entered imputation. After the same post-imputation filters and cross-cohort intersection, 6,955,588 variants remained; post-missingness gave 6,610,616, and HWE $p \geq 1\times10^{-6}$ yielded 6,609,472 variants for analysis. Matched UK Biobank controls carry the same post-imputation set.

***AllofUs case control study.*** From 524 cases with whole genome sequencing (WGS), 32 PCA outliers and 8 related individuals (kinship > 0.0625) were removed; 22 additional samples failed missingness/heterozygosity thresholds, leaving 462 cases. Controls were matched within the AllofUs WGS resource using the same PCAmatchR framework: 1,460 individuals were selected and 74 excluded for missingness/heterozygosity, leaving 1,386 controls. The WGS call set comprised 99,250,816 variants (hg38). Applying the cross-study variant milestones where applicable—autosomal biallelic restriction with MAF ≥ 0.01, post-missingness (< 5 %) and HWE $p \geq 1\times10^{-6}$—and converting hg38→hg19 yielded 15,423,079, 8,130,256, 7,946,982 and finally 7,918,868 variants, respectively. The matching AllofUs controls share the same final variant counts than the cases.

***Meta-analysis and cross-cohort integration.*** Per-study summary statistics (MUSC 1, MUSC 2, All of Us) were harmonized on hg19 and combined using fixed-effects inverse-variance weighting in METAL (March 2020) [7]. We retained autosomal biallelic SNPs present in all three studies, giving 6,249,775 intersection variants. Variants with between-study heterogeneity (Cochran's Q $p \leq 0.01$) were excluded, leaving 6,181,533 high-quality variants in the final meta-analysis. Genome-wide significance was defined as $P < 5\times10^{-8}$. Regional association plots were generated with LocusZoom [8].

### Supplementary Figures and Tables

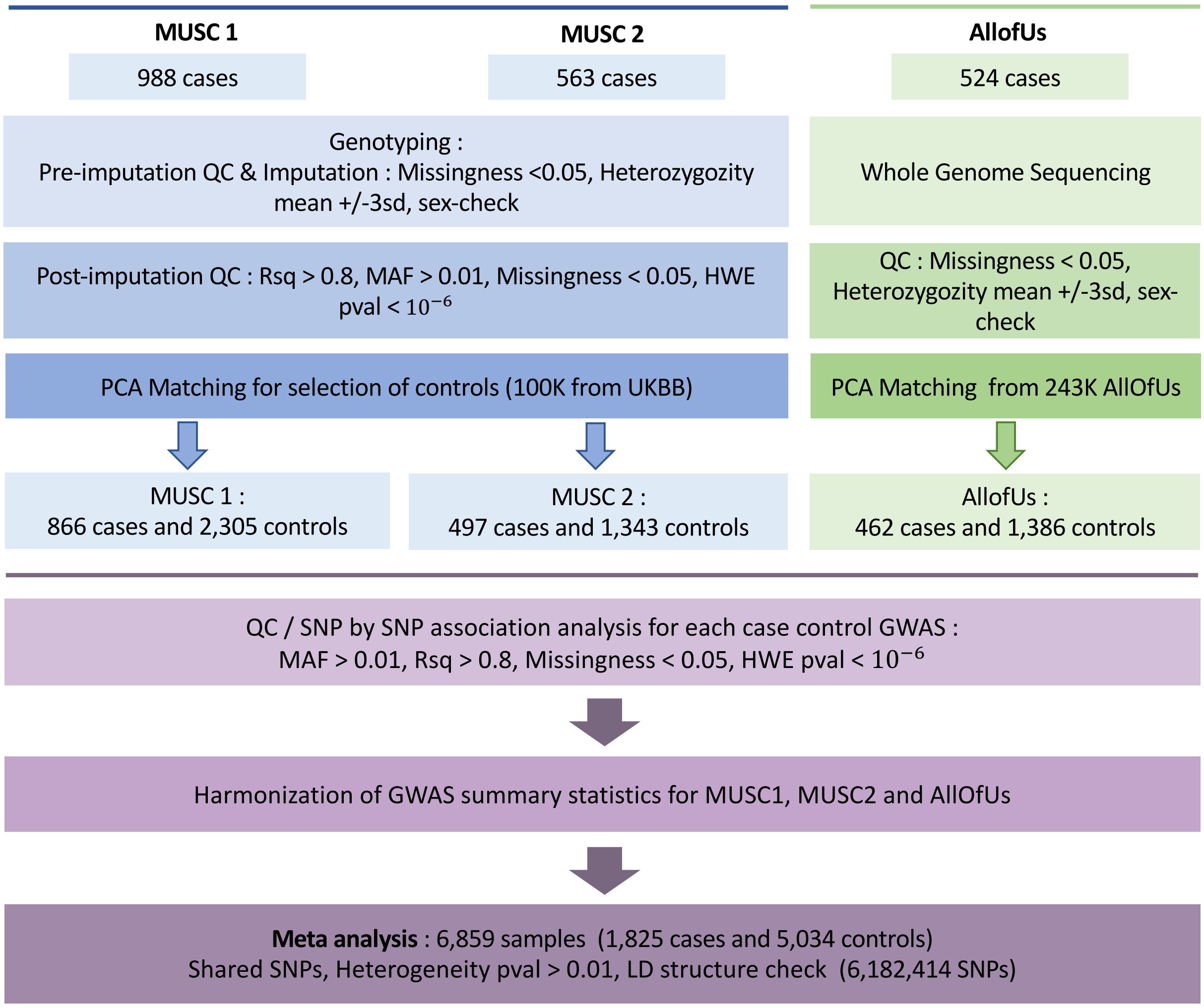


**Supp Figure1. GWAS meta-analysis design.**
Flowchart representing the GWAS meta-analysis design of the three patient cohorts. Cohorts MUSC 1 and MUSC 2 were analyzed together, including the genotyping, the quality control, and the PCA matching steps. The AllOfUs cohort was sequenced, followed by quality control and PCA matching. For all three cohorts, a SNP by SNP association analysis was performed before harmonizing the summary statistics for the meta-analysis.
QC: Quality Control, PCA: Principal Component Analysis, MAF: Minor Allele Frequency, HWE: Hardy–Weinberg Equilibrium.

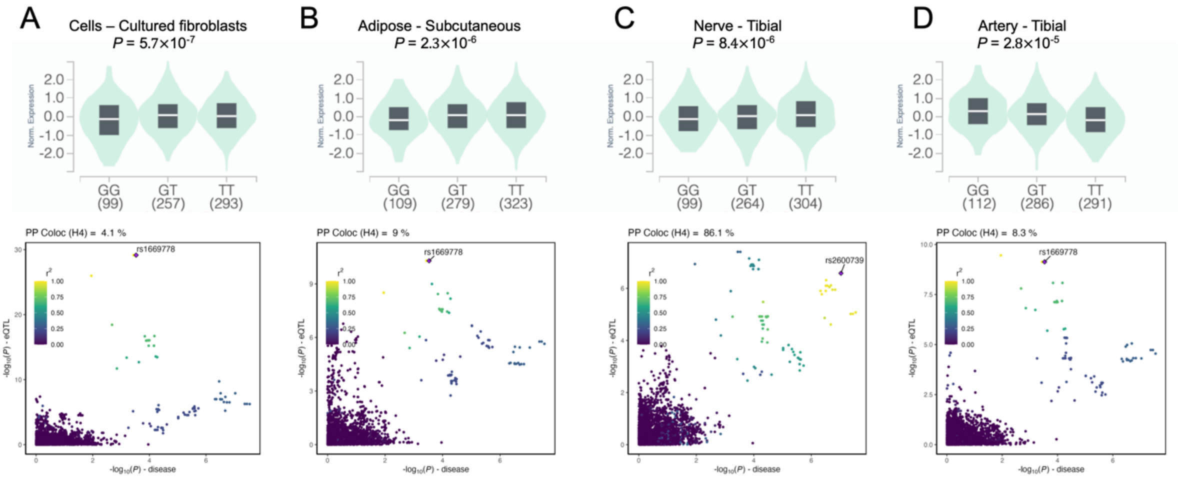


#### Supp Figure 2. eQTL lookup and colocalization at *ACKR3* locus.

GTEx database (v10) was queried with lead SNP at *ACKR3* locus (rs2708184). Significant eQTLs all involved ACKR3 in Cultured fibroblasts (**A**), Adipose tissue (**B**), Tibial Nerve (**C**) and Tibial Artery (**D**). For each eQTL association, violin plot (upper panel) represents the normalized expression of *ACKR3* depending on rs2708184 genotype. Number of individuals for each genotype are indicated in legend. Lower panel represents -log10(*P*-value) of *ACKR3* eQTL association (*y*-axis) vs hEDS association (*x*-axis) for all SNPs within 500kb of lead SNP at *ACKR3* locus. Diamond dot shape indicates the SNP maximizing association to both *ACKR3* eQTL and hEDS. Dot-color indicates linkage desequilibrium (r2) with this SNP in the European subset of 1000G reference panel. Posterior probability for colocalization of both signals is indicated over each graph.

**Supplementary Table1. Clinical descriptive of populations included in the GWAS meta-analysis.**

* Details about ICD-10 codes relatives to potenitally confouding diseases applied to select controls are reported in SuppTable 3.

| **Label** | **MUSC Cases 1 (n=866)** | **UKBB Control Set 1 (n=2305)** | **MUSC Cases 2 (n=497)** | **UKBB Control Set 2 (n=2343)** | **AllofUS (n=526)** | **AllofUS (n=1386)** |
|---|---|---|---|---|---|---|
| **Status** | **Cases** | **Controls** | **Cases** | **Controls** | **Cases** | **Controls** |
| **Type** | Clinical-based | Population-based | Clinical-based | Population-based | Population-based | Population-based |
| **Inclusion Criteria** | Age >12 with a clinical diagnosis of hEDS | None | Age >12 with a clinical diagnosis of hEDS | None | Age >18 (Age >19 in Alabama, Age >21 in Puerto Rico) and WGS data | Age >18 (Age >19 in Alabama, Age >21 in Puerto Rico) and WGS data |
| **Exclusion Criteria** | Age <12 without a clinical diagnosis of hEDS | ICD-Codes confounding diagnosis* | Age <12 without a clinical diagnosis of hEDS | ICD-Codes confounding diagnosis* | Individuals with a diagnosis of hEDS* | Individuals without hEDS, ICD-Codes confounding diagnosis* |
| **Male (n)** | 31 | 1005 | 19 | 664 | 37 | 543 |
| **Female (n)** | 835 | 1300 | 478 | 679 | 425 | 843 |
| **Mean Age (Years) (Std. Dev., Min., Max.)** | 40 (13, 14, 80) | 56 (8,40,70) | 41 (12.7, 13, 76) | 56 (7.9,40,70) | 48 | 61 |

**Supplementary Table 2: Major comorbidities reported in hEDS patients studied.** NA: Not available.

| | MUSC 1 Patients | | MUSC 2 Patients | | | AllofUS hEDS | |
|---|---|---|---|---|---|---|---|
| **Clinical Phenotypes/Traits (Questionnaires)** | N | % | N | % | **Closest item from electronic health records** | N | % |
| Total | 866 | | 497 | | Total | 462 | |
| Joint Dislocations | 533 | 62% | 300 | 60% | Dislocation of joint | 75 | 16% |
| Joint Subluxations | 773 | 89% | 452 | 91% | Subluxation of joint | 24 | 5% |
| Dysautonomia/Postural Orthostatic Tachycardia Syndrome (POTS) | 614 | 71% | 346 | 70% | Disorder of autonomic nervous system | 125 | 27% |
| Mast Cell Activation Syndrome (MCAS) | 362 | 42% | 194 | 39% | Mast cell activation syndrome | <20 | NA |
| Gastrointestinal manifestations (e.g. gastroparesis, IBS, GERD) | 685 | 79% | 408 | 82% | Disorder of digestive system | 402 | 87% |
| Autism spectrum disorder (ASD) | 76 | 9% | 51 | 10% | Autism spectrum disorder | <20 | NA |
| Migraine | 593 | 68% | 341 | 69% | Migraine | 233 | 50% |
| Cranio-cervical instability (CCI)/Atlanto-axial instability (AAI) | 288 | 33% | 166 | 33% | Cervical spine instability | <20 | NA |
| Chiari malformation | 78 | 9% | 42 | 8% | Chiari malformation | <20 | NA |
| Tethered cord syndrome | 64 | 7% | 33 | 7% | Occult spinal dysraphism sequence | <20 | NA |
| Anxiety | 622 | 72% | 379 | 76% | Anxiety | 296 | 64% |
| Depression | 517 | 60% | 311 | 63% | Depressive disorder | 235 | 51% |
| Mitral valve prolapse/regurgitation | 214 | 25% | 109 | 22% | Mitral valve prolapse | 30 | 6% |
| Other heart valve issues (tricuspid, aortic, pulmonic) | 120 | 14% | 69 | 14% | Aortic valve disorder, Tricuspid valve disorder, Pulmonary valve disorder | 42 | 9% |
| Poor wound healing | 528 | 61% | 322 | 65% | Impaired wound healing | <20 | NA |
| Abnormally stretchy skin | 590 | 68% | 351 | 71% | Not Reported | - | NA |
| Abnormal Scarring | 603 | 70% | 361 | 73% | Not Reported | - | NA |
| Myalgic encephalomyelitis/chronic fatigue syndrome (ME/CFS) | 261 | 30% | 161 | 32% | Chronic fatigue syndrome | 89 | 19% |
| Bleeding or clotting problems | 190 | 22% | 115 | 23% | Blood coagulation disorder | 54 | 12% |
| Chronic pain | 763 | 88% | 453 | 91% | Chronic pain | 336 | 73% |
| Abdominal hernia(s) | 185 | 21% | 109 | 22% | Hernia of abdominal cavity | 30 | 6% |
| Pelvic organ prolapse | 165 | 19% | 97 | 20% | Prolapse of female genital organs | 44 | 10% |
| Raynaud's Phenomenon | 393 | 45% | 221 | 44% | Raynaud's disease | NA | NA |

**Supplementary Table 3. List of ICD-codes and their labels applied to exclude controls in the UKBB and AllofUs.**

| ICD-10 Code | Label |
|---|---|
| Q.65-79 | Congenital malformations and deformations of the musculoskeletal system |
| Q.80-89 | Other congenital malformations |
| M.20-25 | Other joint disorders |
| M.30-36 | Systemic connective tissue disorders |
| I.05-09 | Chronic rheumatic heart diseases |
| I.34-37 | Nonrheumatic valve disorders |
| I.42-43 | Cardiomyopathies |
| I.52 | Other heart disorders in diseases classified elsewhere |
| I.71-72 | Aneurysms and dissections |
| I.253-254 | Aneurysm of the heart or coronary arteries |

**Supplementary Table 4 : Per-study sample quanlity control details.**

Counts are shown separately for cases and controls. N start is the initial eligible set (cases: enrolled; controls: available pool before matching, random 100K in the case of UK Biobank). Excl_nonEUR(PCA) = removals as PCA outliers relative to the 1000G European cluster (>6 SD); Excl_related = removals with KING kinship > 0.0625; cells marked "–" are not applicable to UKBB or All of Us. N_after_PCA_match is the subset used for analysis (for controls, the matched subset; for cases, the remainder after pre-imputation ancestry/relatedness QC). Excl_miss/het = removals for missingness (>5 %) or heterozygosity outliers (mean ± 3 SD). Excl_ICD = controls excluded for confounding diagnoses. N_excluded_total is the sum of the exclusion columns listed; for controls it reflects post-match exclusions only. N_GWAS is the final analytic sample size per study/role.

† Missingness/heterozygosity thresholds as above. ‡ PCA relative to 1000G EUR; >6 SD. § KING kinship > 0.0625.

| Case control Study | Status | N start | Excl_nonEUR (PCA)‡ | Excl_related§ | N_after_PCA_match* | Excl_miss/het† | Excl_ICD | N_excluded_total | N_GWAS |
|---|---|---|---|---|---|---|---|---|---|
| MUSC 1 | Cases | 988 | 107 | 12 | 869 | 3 | 0 | 122 | 866 |
| UKBB | Controls | 100,000 | - | - | 2601 | 9 | 287 | 296 | 2305 |
| MUSC 2 | Cases | 563 | 56 | 4 | 503 | 6 | 0 | 66 | 497 |
| UKBB | Controls | 100,000 | - | - | 1506 | 19 | 144 | 163 | 1343 |
| AllOfUs | Cases | 524 | 32 | 8 | 484 | 22 | 0 | 62 | 462 |
| AllOfUs | Controls | 243 | - | - | 1460 | 74 | 0 | 74 | 1386 |

**Supplementary Table 5: Individual case control studies variant-level quanlity control filtering .**

Origin indicates the analysis source: "Genotyped" (MUSC1/MUSC2 cases), "Genotyped+Imputed" (UKBB controls), "WGS" for whole genome sequencing (All of Us). N_raw_variants and the pre-imputation columns (SNVs_nonmonomorphic_only, duplicates_removed, preimp_miss<2%, HRC_align) apply only to genotyped case cohorts; cells marked "–" are not applicable to UKBB or AllofUs. N_source denotes the size of the analysis source (imputed variant set for MUSC/UKBB; WGS callset for All of Us). post_merge_chr1–22(Rsq>0.8;MAF≥0.01) is the autosomal biallelic intersection after imputation quality and MAF filters, consistent with the flow charts. Subsequent filters are post_miss<5%, HWE_p≥1e–6 (evaluated in controls), and liftOver_hg38→hg19 where applicable (All of Us). N_GWAS_variants is the final analytic variant count per study/role. Rows for UKBB and All of Us controls mirror the paired case cohorts after matching to maintain cross-cohort intersection consistency.

| Study | Status | origin | N_raw_variants | Polymorphic SNVs | duplicates_ removed | preimp_miss (<2%) | HRC_align | N_source (imputed/WGS) | post_merge_chr1-22 (Rsq>0.8;MAF≥0.01) | post_miss (<5%) | HWE (p≥1e-6) | liftOver hg38→hg19 | SNVs in GWAS |
|---|---|---|---|---|---|---|---|---|---|---|---|---|---|
| MUSC 1 | Cases | Genotyped | 730,059 | 599,610 | 595,621 | 576,098 | 551,645 | 40,355,712 | 7,021,115 | 6,686,384 | 6,685,653 | - | 6,685,653 |
| UKBB | Controls | Genotyped+Imputed | 93,095,623 | - | - | - | - | 93,095,623 | 7,021,115 | 6,686,384 | 6,685,653 | - | 6,685,653 |
| MUSC 2 | Cases | Genotyped | 730,059 | 587,181 | 583,354 | 553,424 | 531,821 | 40,355,712 | 6,955,588 | 6,610,616 | 6,609,472 | - | 6,609,472 |
| UKBB | Controls | Genotyped+Imputed | 93,095,623 | - | - | - | - | 93,095,623 | 6,955,588 | 6,610,616 | 6,609,472 | - | 6,609,472 |
| AllOfUs | Cases | WGS | 99,250,816 | - | - | - | - | 99,250,816 | 15,423,079 | 8,130,256 | 7,946,982 | 7,918,868 | 7,918,868 |
| AllOfUs | Controls | WGS | 99,250,816 | - | - | - | - | 99,250,816 | 15,423,079 | 8,130,256 | 7,946,982 | 7,918,868 | 7,918,868 |

**Supplementary Table 6**: Epigenomic datasets used for annotation. Datasets were used as bigWig files.

| Cluster/tissue | Epigenomic Mark | ID | Source |
|---|---|---|---|
| Pancreatic Acinar Cell | ATAC (single nuclei) | Acinar | Human Enhancer Atlas |
| Adipocyte | ATAC (single nuclei) | Adipocyte | Human Enhancer Atlas |
| Cardiac Fibroblasts | ATAC (single nuclei) | Cardiac Fibroblast | Human Enhancer Atlas |
| Colon Epithelial Cell 1 | ATAC (single nuclei) | Colon Epithelial 1 | Human Enhancer Atlas |
| Cortical Epithelial-like | ATAC (single nuclei) | Cortical Epithelial | Human Enhancer Atlas |
| Endothelial Cell (General) 1 | ATAC (single nuclei) | Endothelial General 1 | Human Enhancer Atlas |
| Endothelial Cell (General) 2 | ATAC (single nuclei) | Endothelial General 2 | Human Enhancer Atlas |
| Esophageal Epithelial Cell | ATAC (single nuclei) | Esophageal Epithelial | Human Enhancer Atlas |
| Zona Fasciculata Cortical Cell | ATAC (single nuclei) | Fasciculata | Human Enhancer Atlas |
| Fibroblast (Epithelial) | ATAC (single nuclei) | Fibro Epithelial | Human Enhancer Atlas |
| Fibroblast (Gastrointestinal) | ATAC (single nuclei) | Fibro GI | Human Enhancer Atlas |
| Fibroblast (General) | ATAC (single nuclei) | Fibro General | Human Enhancer Atlas |
| Fibroblast (Liver Adrenal) | ATAC (single nuclei) | Fibro Liver Adrenal | Human Enhancer Atlas |
| Fibroblast (Sk Muscle Associated) | ATAC (single nuclei) | Fibro Muscle | Human Enhancer Atlas |
| Fibroblast (Peripheral Nerve) | ATAC (single nuclei) | Fibro Nerve | Human Enhancer Atlas |
| Thyroid Follicular Cell | ATAC (single nuclei) | Follicular | Human Enhancer Atlas |
| Zona Glomerulosa Cortical Cell | ATAC (single nuclei) | Glomerulosa | Human Enhancer Atlas |
| Hepatocyte | ATAC (single nuclei) | Hepatocyte | Human Enhancer Atlas |
| Keratinocyte 1 | ATAC (single nuclei) | Keratinocyte 1 | Human Enhancer Atlas |
| Macrophage (General) | ATAC (single nuclei) | Macrophage General | Human Enhancer Atlas |
| Mast Cell | ATAC (single nuclei) | Mast | Human Enhancer Atlas |
| Melanocyte | ATAC (single nuclei) | Melanocyte | Human Enhancer Atlas |
| Mesothelial Cell | ATAC (single nuclei) | Mesothelial | Human Enhancer Atlas |
| Myoepithelial (Skin) | ATAC (single nuclei) | Myoepithelial | Human Enhancer Atlas |
| Peripheral Nerve Stromal | ATAC (single nuclei) | Nerve Stromal | Human Enhancer Atlas |
| Schwann Cell (General) | ATAC (single nuclei) | Schwann General | Human Enhancer Atlas |
| Basal Epidermal (Skin) | ATAC (single nuclei) | Skin Basal Epidermal | Human Enhancer Atlas |
| Eccrine Epidermal (Skin) | ATAC (single nuclei) | Skin Eccrine Epidermal | Human Enhancer Atlas |
| Granular Epidermal (Skin) | ATAC (single nuclei) | Skin Granular Epidermal | Human Enhancer Atlas |
| Smooth Muscle (Colon) 1 | ATAC (single nuclei) | Sm Ms Colon 1 | Human Enhancer Atlas |
| Smooth Muscle (General) | ATAC (single nuclei) | Sm Ms General | Human Enhancer Atlas |
| Smooth Muscle (Esophageal Muscularis) 1 | ATAC (single nuclei) | Sm Ms Muscularis 1 | Human Enhancer Atlas |
| T Lymphocyte 1 (CD8+) | ATAC (single nuclei) | T Lymphocyte 1 (CD8+) | Human Enhancer Atlas |
| T lymphocyte 2 (CD4+) | ATAC (single nuclei) | T lymphocyte 2 (CD4+) | Human Enhancer Atlas |
| Transitional Zone Cortical Cell | ATAC (single nuclei) | Transitional Cortical | Human Enhancer Atlas |
| Type I Skeletal Myocyte | ATAC (single nuclei) | Type I Skeletal Myocyte | Human Enhancer Atlas |
| Type II Skeletal Myocyte | ATAC (single nuclei) | Type II Skeletal Myocyte | Human Enhancer Atlas |
| Ventricular Cardiomyocyte | ATAC (single nuclei) | V Cardiomyocyte | Human Enhancer Atlas |
| Vascular Smooth Muscle 1 | ATAC (single nuclei) | Vasc Sm Muscle 1 | Human Enhancer Atlas |
| Vascular Smooth Muscle 2 | ATAC (single nuclei) | Vasc Sm Muscle 2 | Human Enhancer Atlas |
| PVALB+ GABAergic neurons | ATAC (single nuclei) | PVALB-1 | Human Brain enhancer atlas |
| SST+ GABAergic neurons | ATAC (single nuclei) | SST-1 | Human Brain enhancer atlas |
| VIP+ GABAergic neurons | ATAC (single nuclei) | VIP-1 | Human Brain enhancer atlas |
| Granule neurons from cerebellum | ATAC (single nuclei) | CBGRC | Human Brain enhancer atlas |
| Cholinergic neurons | ATAC (single nuclei) | CHO | Human Brain enhancer atlas |
| L6 corticothalamic (CT) projection neurons | ATAC (single nuclei) | CT-1 | Human Brain enhancer atlas |
| Intratelencephalic projecting neurons, cortical l | ATAC (single nuclei) | IT-L2/3-1 | Human Brain enhancer atlas |
| Intratelencephalic projecting neurons, cortical l | ATAC (single nuclei) | IT-L2/3-2 | Human Brain enhancer atlas |
| Intratelencephalic projecting neurons, cortical l | ATAC (single nuclei) | IT-L2/3-3 | Human Brain enhancer atlas |
| Non-telencephalon astrocytes | ATAC (single nuclei) | ASCNT-1 | Human Brain enhancer atlas |
| Telencephalon astrocytes | ATAC (single nuclei) | ASCT-1 | Human Brain enhancer atlas |
| Microglia | ATAC (single nuclei) | MGC-1 | Human Brain enhancer atlas |
| Microglia | ATAC (single nuclei) | MGC-2 | Human Brain enhancer atlas |
| Oligodendrocytes | ATAC (single nuclei) | OGC-1 | Human Brain enhancer atlas |
| Oligodendrocytes | ATAC (single nuclei) | OGC-2 | Human Brain enhancer atlas |
| Oligodendrocytes | ATAC (single nuclei) | OGC-3 | Human Brain enhancer atlas |
| Adipose | ATAC | ENCFF094EYJ | ENCODE |
| Adipose | H3K27ac | ENCFF608YXU | ENCODE |
| Adrenal | H3K4me3 | ENCFF053KMZ | ENCODE |

| | | | |
|---|---|---|---|
| Adrenal | H3K4me3 | ENCFF700TZZ | ENCODE |
| Adrenal | ATAC | ENCFF083NYF | ENCODE |
| Adrenal | ATAC | ENCFF854MVA | ENCODE |
| Adrenal | H3K27ac | ENCFF860MMV | ENCODE |
| Adrenal | H3K27ac | ENCFF464QEK | ENCODE |
| Colon | H3K27ac | ENCFF322NLT | ENCODE |
| Colon | H3K27ac | ENCFF468UEP | ENCODE |
| Coronary artery | H3K27ac | ENCFF130NUG | ENCODE |
| Coronary artery | H3K27ac | ENCFF476MBG | ENCODE |
| Coronary artery | H3K4me3 | ENCFF811RQX | ENCODE |
| Heart LV | ATAC | ENCFF566TSW | ENCODE |
| Heart LV | ATAC | ENCFF065BYP | ENCODE |
| Heart LV | H3K27ac | ENCFF718XVH | ENCODE |
| Heart LV | H3K4me1 | ENCFF767RQM | ENCODE |
| Heart LV | H3K4me3 | ENCFF108LHZ | ENCODE |
| Heart RAAR | H3K27ac | ENCFF998CAS | ENCODE |
| Heart RAAR | H3K4me1 | ENCFF663AHY | ENCODE |
| Heart RAAR | H3K4me3 | ENCFF956EDO | ENCODE |
| Liver | ATAC | ENCFF341RHY | ENCODE |
| Liver | H3K27ac | ENCFF555QGS | ENCODE |
| Liver | H3K4me1 | ENCFF702XFW | ENCODE |
| Liver | H3K4me3 | ENCFF215LWY | ENCODE |
| Pancreas | ATAC | ENCFF246NCJ | ENCODE |
| Pancreas | H3K27ac | ENCFF474LLN | ENCODE |
| Pancreas | H3K4me1 | ENCFF198TAL | ENCODE |
| Pancreas | H3K4me3 | ENCFF324TNC | ENCODE |
| Psoas | Dnase-seq | ENCFF172AJV | ENCODE |
| Psoas | H3K27ac | ENCFF184XXA | ENCODE |
| Psoas | H3K27ac | ENCFF378PPV | ENCODE |
| Skin | Dnase-seq | ENCFF548VEV | ENCODE |
| Skin | Dnase-seq | ENCFF642VOM | ENCODE |
| Skin | H3K27ac | ENCFF197ZLE | ENCODE |
| Skin | H3K27ac | ENCFF702ZYG | ENCODE |
| Skin | H3K27ac | ENCFF868JPL | ENCODE |
| Skin | H3K4me1 | ENCFF436HQD | ENCODE |
| Skin | H3K4me3 | ENCFF697CGX | ENCODE |
| Thyroid | ATAC | ENCFF284QOX | ENCODE |
| Thyroid | H3K27ac | ENCFF578GZL | ENCODE |
| Thyroid | H3K4me1 | ENCFF687HLG | ENCODE |
| Thyroid | H3K4me3 | ENCFF771GOG | ENCODE |
| Tibial Nerve | ATAC | ENCFF526USD | ENCODE |
| Tibial Nerve | H3K27ac | ENCFF574FEG | ENCODE |

**Supplementary Table 7. GWAS summary statistics used for LD-score regression genetic correlation analyses.**

The table lists, for each analysed phenotype, the descriptive label (phenotype), the internal code used throughout figures (Abreviation), category, sample counts Neff (computed as 4/(1/Ncases+1/Nctrl), the source/consortium, the article reference, and the download link (GWAS Catalog harmonised files when available). All summary statistics were aligned to the same effect allele on hg19/GRCh37, restricted to HapMap3 SNPs, and reformatted with munge_sumstats.py. Analyses were limited to European-ancestry datasets; per-trait cohort composition and final Ns are provided here. Phenotypes include Myalgic encephalomyelitis/chronic fatigue syndrome, autonomic nervous system disorders, chronic pain, joint hypermobility, depression, autism spectrum disorder, anxiety, migraine, irritable bowel syndrome, gastroesophageal reflux disease, gastroparesis, inguinal/umbilical/ventral/abdominal hernia, mitral valve prolapse, tricuspid valve disease, and pelvic organ prolapse . Note: hernia subtypes are listed separately in the table but counted as one non-redundant phenotype family in multiple-testing procedures (see Methods).

| Abreviation | Phenotype | Category | N_cases | N_ctrls | Neff | Source | From | Reference | link |
|---|---|---|---|---|---|---|---|---|---|
| ME/CFS | Myalgic encephalomyelitis / chronic fatigue syndrome | Pain & Autonomic | 15579 | 259909 | 58792 | DECODE | DECODE | https://www.pure.ed.ac.uk/ws/portalfiles/portal/533352490/Preprint.pdf | https://osf.io/rgqs3/files/osfstorage/6893a8af46cceef0f3b218db |
| ANS | Dysautonomia (autonomic nervous system) | Pain & Autonomic | 14559 | 421084 | 56290 | GWAS Catalog | MVP | PMID: 39024449 | https://ftp.ebi.ac.uk/pub/databases/gwas/summary_statistics/GCST90475001-GCST90476000/GCST90475834/harmonised/GCST90475834.h.tsv.gz |
| CP | Chronic pain | Pain & Autonomic | 54185 | 354536 | 188006 | GWAS Catalog | MVP | PMID: 39024449 | https://ftp.ebi.ac.uk/pub/databases/gwas/summary_statistics/GCST90477001-GCST90478000/GCST90477533/harmonised/GCST90477533.h.tsv.gz |
| FIB | Fibromyaglia | Pain & Autonomic | 2149 | 433822 | 8554 | GWAS Catalog | UKBB | PMID: 37219871 | https://ftp.ebi.ac.uk/pub/databases/gwas/summary_statistics/GCST90129001-GCST90130000/GCST90129439/harmonised/GCST90129439.h.tsv.gz |
| JH | Joint hypermobility (code : M13) | HEDS Spectrum | 2150 | 484260 | 8562 | FinnGen | FinnGen (R12) | finngen | https://storage.googleapis.com/finngen-public-data-r12/summary_stats/release/finngen_R12_M13_HYPERMOBILITY.gz |
| MDD | Depression | Psychiatric/Neurological | 410965 | 1583375 | 1305117 | PGC | PGC | PMID: 39814019 | https://pgc.unc.edu/for-researchers/download-results/ |
| ASD | Autism spectrum disorder | Psychiatric/Neurological | 18382 | 27969 | 44368 | PGC | IPSYCH | PMID: 30804558 | https://pgc.unc.edu/for-researchers/download-results/ |
| ANX | Anxiety | Psychiatric/Neurological | 7016 | 14745 | 19016 | PGC | PGC | PMID: 26754954 | https://pgc.unc.edu/for-researchers/download-results/ |
| MIG | Migraine | Psychiatric/Neurological | 31836 | 405831 | 118081 | GWAS Catalog | MVP | PMID: 39024449 | https://ftp.ebi.ac.uk/pub/databases/gwas/summary_statistics/GCST90475001-GCST90476000/GCST90475837/harmonised/GCST90475837.h.tsv.gz |
| IBS | Iirritable bowel syndrome | Gastrointestinal manifestations | 53400 | 433201 | 190159 | GWAS Catalog | Meta-analysis | PMID: 34741163 | https://ftp.ebi.ac.uk/pub/databases/gwas/summary_statistics/GCST90016001-GCST90017000/GCST90016564/harmonised/34741163-GCST90016564-EFO_0000555.h.tsv.gz |
| GERD | Gastroesophageal reflux disease | Gastrointestinal manifestations | 175991 | 236737 | 403787 | GWAS Catalog | MVP | PMID: 39024449 | https://ftp.ebi.ac.uk/pub/databases/gwas/summary_statistics/GCST90476001-GCST90477000/GCST90476047/harmonised/GCST90476047.h.tsv.gz |
| GAST | Gastroparesis | Gastrointestinal manifestations | 2813 | 446383 | 11182 | GWAS Catalog | MVP | PMID: 39024449 | https://ftp.ebi.ac.uk/pub/databases/gwas/summary_statistics/GCST90478001-GCST90479000/GCST90478363/harmonised/GCST90478363.h.tsv.gz |
| AH | Abdominal hernia | Hernias | 71035 | 345335 | 235664 | GWAS Catalog | MVP | PMID: 39024449 | https://ftp.ebi.ac.uk/pub/databases/gwas/summary_statistics/GCST90476001-GCST90477000/GCST90476059/harmonised/GCST90476059.h.tsv.gz |
| VH | Ventral hernia | Hernias | 9512 | 433175 | 37230 | GWAS Catalog | MVP | PMID: 39024449 | https://ftp.ebi.ac.uk/pub/databases/gwas/summary_statistics/GCST90476001-GCST90477000/GCST90476063/harmonised/GCST90476063.h.tsv.gz |
| IH | Inguinal hernia | Hernias | 30294 | 410088 | 112840 | GWAS Catalog | MVP | PMID: 39024449 | https://ftp.ebi.ac.uk/pub/databases/gwas/summary_statistics/GCST90476001-GCST90477000/GCST90476060/harmonised/GCST90476060.h.tsv.gz |
| UH | Umbilical hernia | Hernias | 15292 | 428443 | 59060 | GWAS Catalog | MVP | PMID: 39024449 | https://ftp.ebi.ac.uk/pub/databases/gwas/summary_statistics/GCST90476001-GCST90477000/GCST90476062/harmonised/GCST90476062.h.tsv.gz |
| MVP | Mitral valve prolapse/regurgitation | Circulatory | 4884 | 434649 | 19319 | Meta-Analysis | Roselli et al | PMID:35245370 | https://cvd.hugeamp.org/dinspector.html?dataset=Roselli2022_MVP_EU |
| TrVD | Other heart valve issues (tricuspid) | Circulatory | 1353 | 443776 | 5396 | GWAS Catalog | MVP | PMID: 39024449 | https://ftp.ebi.ac.uk/pub/databases/gwas/summary_statistics/GCST90477001-GCST90478000/GCST90477835/harmonised/GCST90477835.h.tsv.gz |
| POP | Pelvic organ prolapse | Other | 11966 | 396730 | 46463 | GWAS Catalog | UKBB | PMID: 30104761 | https://ftp.ebi.ac.uk/pub/databases/gwas/summary_statistics/GCST90436001-GCST90437000/GCST90436496/GCST90436496.tsv.gz |

**Supp Table 8. Lead SNPs in two hEDS loci meeting genome-wide suggestive significance ($P<1\times10^{-4}$) across meta-analysis and individual cohorts**

This table summarizes the association results for all SNPs identified at the two genome-wide significant loci (lead SNP +/- 500 kb region) reaching a suggestive significance threshold of $P<1\times10^{-4}$ in the hypermobile Ehlers-Danlos Syndrome (hEDS) GWAS meta-analysis . We report the effect allele (EA), other allele (OA) and effect allele frequency (EAF), odds ratio with 95% confidence interval [OR (95% CI)], p-value (P) are provided for the individual case control studies: MUSC 1, MUSC2, and AllOfUs. Direction of effect and heterogeneity p-value (Het P) from the meta-analysis are also reported. Abbreviations: hEDS: hypermobile Ehlers-Danlos Syndrome, EAF: effect allele frequency, OR: odds

| Variant | | | | | hEDS GWAS meta-analysis | | | | | hEDS GWAS MUSC 1 | | | hEDS GWAS MUSC2 | | | hEDS GWAS AllOfUs | | |
|---|---|---|---|---|---|---|---|---|---|---|---|---|---|---|---|---|---|---|
| Chr | Position | rsID | EA | OA | EAF | OR (95% CI) | P | Direction | Het P | EAF | OR (95% CI) | P | EAF | OR (95% CI) | P | EAF | OR (95% CI) | P |
| 2 | 237545175 | rs2600767 | A | G | 0.12 | 1.31 (1.15 ; 1.48) | 4.44E-05 | +++ | 0.16 | 0.12 | 1.32 (1.09 ; 1.60) | 4.36E-03 | 0.12 | 1.55 (1.20 ; 2.0) | 7.31E-04 | 0.12 | 1.11 (0.88 ; 1.40) | 3.82E-01 |
| 2 | 237545441 | rs2720149 | T | G | 0.25 | 1.25 (1.14 ; 1.38) | 7.21E-06 | +++ | 0.89 | 0.24 | 1.27 (1.10 ; 1.47) | 1.40E-03 | 0.25 | 1.28 (1.05 ; 1.56) | 1.36E-02 | 0.24 | 1.21 (1.01 ; 1.45) | 4.34E-02 |
| 2 | 237545512 | rs2464850 | A | C | 0.25 | 1.26 (1.14 ; 1.39) | 4.24E-06 | +++ | 0.85 | 0.25 | 1.27 (1.10 ; 1.47) | 1.13E-03 | 0.25 | 1.30 (1.07 ; 1.58) | 9.26E-03 | 0.24 | 1.21 (1.01 ; 1.45) | 4.28E-02 |
| 2 | 237545560 | rs2443941 | T | C | 0.75 | 0.79 (0.72 ; 0.88) | 4.19E-06 | --- | 0.84 | 0.75 | 0.78 (0.68 ; 0.90) | 8.97E-04 | 0.75 | 0.78 (0.64 ; 0.94) | 1.08E-02 | 0.76 | 0.83 (0.69 ; 1.00) | 4.59E-02 |
| 2 | 237545629 | rs2708182 | A | G | 0.75 | 0.79 (0.72 ; 0.88) | 4.18E-06 | --- | 0.85 | 0.75 | 0.79 (0.68 ; 0.91) | 1.12E-03 | 0.75 | 0.77 (0.63 ; 0.94) | 9.26E-03 | 0.76 | 0.83 (0.69 ; 0.99) | 4.28E-02 |
| 2 | 237545680 | rs2708181 | A | G | 0.75 | 0.80 (0.72 ; 0.88) | 7.20E-06 | --- | 0.89 | 0.76 | 0.79 (0.68 ; 0.91) | 1.41E-03 | 0.75 | 0.78 (0.64 ; 0.95) | 1.35E-02 | 0.76 | 0.83 (0.69 ; 0.99) | 4.34E-02 |
| 2 | 237546408 | rs1796436 | A | G | 0.76 | 0.79 (0.71 ; 0.87) | 1.88E-06 | --- | 0.74 | 0.76 | 0.77 (0.66 ; 0.89) | 3.59E-04 | 0.75 | 0.77 (0.63 ; 0.94) | 8.50E-03 | 0.76 | 0.84 (0.70 ; 1.00) | 5.56E-02 |
| 2 | 237546678 | rs1796437 | T | C | 0.70 | 0.83 (0.76 ; 0.91) | 8.46E-05 | --- | 0.77 | 0.70 | 0.83 (0.72 ; 0.95) | 7.40E-03 | 0.70 | 0.79 (0.65 ; 0.95) | 1.31E-02 | 0.70 | 0.87 (0.73 ; 1.03) | 1.03E-01 |
| 2 | 237546838 | rs1796439 | A | G | 0.76 | 0.79 (0.71 ; 0.87) | 1.89E-06 | --- | 0.74 | 0.76 | 0.77 (0.66 ; 0.89) | 3.59E-04 | 0.75 | 0.77 (0.63 ; 0.94) | 8.53E-03 | 0.76 | 0.84 (0.70 ; 1.00) | 5.56E-02 |
| 2 | 237547016 | rs1796440 | A | G | 0.76 | 0.79 (0.72 ; 0.87) | 3.46E-06 | --- | 0.78 | 0.76 | 0.77 (0.67 ; 0.89) | 4.79E-04 | 0.75 | 0.78 (0.64 ; 0.95) | 1.30E-02 | 0.76 | 0.84 (0.70 ; 1.00) | 5.55E-02 |
| 2 | 237547067 | rs1620042 | A | G | 0.25 | 1.27 (1.15 ; 1.4) | 1.72E-06 | +++ | 0.73 | 0.24 | 1.31 (1.13 ; 1.51) | 3.29E-04 | 0.25 | 1.30 (1.07 ; 1.58) | 8.35E-03 | 0.24 | 1.19 (1.00 ; 1.43) | 5.56E-02 |
| 2 | 237547072 | rs1796442 | T | G | 0.25 | 1.27 (1.15 ; 1.4) | 1.72E-06 | +++ | 0.73 | 0.24 | 1.30 (1.13 ; 1.51) | 3.29E-04 | 0.25 | 1.30 (1.07 ; 1.58) | 8.34E-03 | 0.24 | 1.19 (1.00 ; 1.43) | 5.56E-02 |
| 2 | 237547583 | rs2600766 | T | G | 0.24 | 1.27 (1.15 ; 1.4) | 2.19E-06 | +++ | 0.76 | 0.24 | 1.30 (1.12 ; 1.50) | 4.47E-04 | 0.25 | 1.30 (1.07 ; 1.58) | 8.59E-03 | 0.24 | 1.20 (1.00 ; 1.43) | 5.31E-02 |
| 2 | 237547598 | rs2600765 | A | G | 0.76 | 0.79 (0.71 ; 0.87) | 2.13E-06 | --- | 0.79 | 0.76 | 0.77 (0.67 ; 0.89) | 4.98E-04 | 0.75 | 0.77 (0.63 ; 0.94) | 8.58E-03 | 0.76 | 0.83 (0.69 ; 1.00) | 4.76E-02 |
| 2 | 237547638 | rs2600764 | T | G | 0.70 | 0.83 (0.76 ; 0.91) | 9.49E-05 | --- | 0.78 | 0.70 | 0.83 (0.73 ; 0.96) | 8.76E-03 | 0.70 | 0.79 (0.65 ; 0.95) | 1.33E-02 | 0.70 | 0.86 (0.73 ; 1.03) | 9.78E-02 |
| 2 | 237547837 | rs2600763 | T | C | 0.12 | 1.30 (1.15 ; 1.48) | 4.90E-05 | +++ | 0.19 | 0.12 | 1.33 (1.10 ; 1.61) | 3.74E-03 | 0.12 | 1.53 (1.19 ; 1.98) | 1.08E-03 | 0.12 | 1.11 (0.88 ; 1.40) | 3.81E-01 |
| 2 | 237547860 | rs2245594 | T | G | 0.24 | 1.26 (1.14 ; 1.39) | 4.55E-06 | +++ | 0.79 | 0.24 | 1.29 (1.12 ; 1.50) | 5.25E-04 | 0.25 | 1.27 (1.05 ; 1.55) | 1.61E-02 | 0.24 | 1.20 (1.00 ; 1.43) | 5.57E-02 |
| 2 | 237547863 | rs2245593 | A | G | 0.76 | 0.79 (0.71 ; 0.87) | 1.95E-06 | --- | 0.74 | 0.76 | 0.77 (0.66 ; 0.89) | 3.48E-04 | 0.76 | 0.77 (0.63 ; 0.94) | 9.34E-03 | 0.76 | 0.84 (0.70 ; 1.00) | 5.50E-02 |
| 2 | 237549355 | rs2720151 | T | G | 0.75 | 0.79 (0.72 ; 0.87) | 2.84E-06 | --- | 0.74 | 0.75 | 0.77 (0.67 ; 0.89) | 4.97E-04 | 0.75 | 0.77 (0.63 ; 0.94) | 8.70E-03 | 0.76 | 0.84 (0.70 ; 1.01) | 6.11E-02 |
| 2 | 237549430 | rs2245481 | T | G | 0.75 | 0.79 (0.71 ; 0.87) | 1.38E-06 | --- | 0.75 | 0.75 | 0.77 (0.67 ; 0.89) | 4.22E-04 | 0.75 | 0.76 (0.62 ; 0.92) | 5.88E-03 | 0.76 | 0.83 (0.69 ; 1.00) | 4.88E-02 |
| 2 | 237549819 | rs2720152 | A | G | 0.25 | 1.27 (1.15 ; 1.4) | 1.88E-06 | +++ | 0.71 | 0.25 | 1.30 (1.12 ; 1.50) | 4.48E-04 | 0.25 | 1.32 (1.08 ; 1.6) | 5.95E-03 | 0.24 | 1.19 (0.99 ; 1.43) | 6.02E-02 |
| 2 | 237549850 | rs2600753 | A | G | 0.25 | 1.26 (1.15 ; 1.39) | 3.13E-06 | +++ | 0.74 | 0.25 | 1.29 (1.12 ; 1.50) | 5.38E-04 | 0.25 | 1.30 (1.07 ; 1.58) | 8.79E-03 | 0.24 | 1.19 (0.99 ; 1.43) | 6.20E-02 |
| 2 | 237550079 | rs2720153 | C | G | 0.24 | 1.27 (1.15 ; 1.4) | 1.76E-06 | +++ | 0.78 | 0.24 | 1.31 (1.14 ; 1.52) | 2.49E-04 | 0.25 | 1.27 (1.05 ; 1.55) | 1.60E-02 | 0.24 | 1.21 (1.01 ; 1.45) | 4.23E-02 |
| 2 | 237550197 | rs2952665 | A | G | 0.70 | 0.82 (0.74 ; 0.9) | 2.30E-05 | --- | 0.75 | 0.70 | 0.81 (0.71 ; 0.93) | 2.94E-03 | 0.70 | 0.78 (0.64 ; 0.94) | 9.79E-03 | 0.71 | 0.86 (0.72 ; 1.02) | 8.46E-02 |
| 2 | 237551637 | rs2600747 | T | C | 0.81 | 0.80 (0.72 ; 0.89) | 4.90E-05 | --- | 0.66 | 0.80 | 0.82 (0.70 ; 0.96) | 1.28E-02 | 0.81 | 0.74 (0.60 ; 0.91) | 5.20E-03 | 0.81 | 0.84 (0.69 ; 1.01) | 6.90E-02 |
| 2 | 237551662 | rs2600746 | C | G | 0.33 | 1.29 (1.18 ; 1.41) | 3.00E-08 | +++ | 0.87 | 0.33 | 1.32 (1.16 ; 1.51) | 3.80E-05 | 0.33 | 1.25 (1.04 ; 1.5) | 1.70E-02 | 0.32 | 1.28 (1.08 ; 1.51) | 3.92E-03 |
| 2 | 237552327 | rs2708184 | T | G | 0.67 | 0.77 (0.71 ; 0.85) | 2.32E-08 | --- | 0.83 | 0.67 | 0.75 (0.66 ; 0.86) | 2.47E-05 | 0.67 | 0.80 (0.67 ; 0.96) | 1.70E-02 | 0.68 | 0.79 (0.67 ; 0.93) | 4.46E-03 |
| 2 | 237552810 | rs1669777 | T | C | 0.33 | 1.29 (1.18 ; 1.41) | 2.51E-08 | +++ | 0.87 | 0.33 | 1.33 (1.16 ; 1.51) | 3.37E-05 | 0.33 | 1.25 (1.04 ; 1.5) | 1.59E-02 | 0.32 | 1.28 (1.08 ; 1.51) | 3.92E-03 |
| 2 | 237553041 | rs1669776 | T | C | 0.19 | 1.24 (1.11 ; 1.38) | 7.36E-05 | +++ | 0.72 | 0.20 | 1.22 (1.04 ; 1.43) | 1.22E-02 | 0.19 | 1.33 (1.08 ; 1.65) | 8.05E-03 | 0.19 | 1.19 (0.98 ; 1.44) | 7.95E-02 |
| 2 | 237553220 | rs1669775 | T | C | 0.81 | 0.81 (0.73 ; 0.9) | 9.59E-05 | --- | 0.69 | 0.80 | 0.82 (0.70 ; 0.96) | 1.36E-02 | 0.81 | 0.75 (0.61 ; 0.93) | 8.07E-03 | 0.81 | 0.85 (0.70 ; 1.03) | 9.18E-02 |
| 2 | 237553413 | rs2708183 | A | G | 0.81 | 0.81 (0.73 ; 0.9) | 7.86E-05 | --- | 0.64 | 0.80 | 0.82 (0.70 ; 0.96) | 1.32E-02 | 0.81 | 0.74 (0.60 ; 0.92) | 6.12E-03 | 0.81 | 0.85 (0.70 ; 1.03) | 9.33E-02 |
| 2 | 237554565 | rs2708188 | C | G | 0.19 | 1.24 (1.11 ; 1.37) | 9.06E-05 | +++ | 0.71 | 0.20 | 1.22 (1.05 ; 1.43) | 1.20E-02 | 0.19 | 1.33 (1.07 ; 1.65) | 8.84E-03 | 0.19 | 1.18 (0.97 ; 1.43) | 9.18E-02 |
| 2 | 237554723 | rs2708186 | A | T | 0.81 | 0.80 (0.72 ; 0.89) | 5.13E-05 | --- | 0.63 | 0.80 | 0.82 (0.70 ; 0.96) | 1.18E-02 | 0.81 | 0.74 (0.59 ; 0.91) | 4.85E-03 | 0.81 | 0.84 (0.69 ; 1.02) | 8.10E-02 |
| 2 | 237554988 | rs1796444 | A | G | 0.81 | 0.81 (0.73 ; 0.9) | 8.07E-05 | --- | 0.67 | 0.80 | 0.82 (0.70 ; 0.96) | 1.25E-02 | 0.81 | 0.75 (0.60 ; 0.92) | 7.11E-03 | 0.81 | 0.85 (0.70 ; 1.03) | 9.18E-02 |
| 2 | 237555696 | rs2600744 | A | G | 0.81 | 0.80 (0.72 ; 0.89) | 5.06E-05 | --- | 0.63 | 0.80 | 0.82 (0.70 ; 0.96) | 1.17E-02 | 0.81 | 0.74 (0.59 ; 0.91) | 4.85E-03 | 0.81 | 0.84 (0.69 ; 1.02) | 8.10E-02 |
| 2 | 237555822 | rs1631825 | A | T | 0.81 | 0.81 (0.73 ; 0.9) | 7.84E-05 | --- | 0.57 | 0.80 | 0.82 (0.70 ; 0.96) | 1.15E-02 | 0.81 | 0.74 (0.59 ; 0.91) | 4.84E-03 | 0.81 | 0.86 (0.71 ; 1.04) | 1.22E-01 |
| 2 | 237556105 | rs2600743 | T | C | 0.81 | 0.81 (0.73 ; 0.9) | 8.39E-05 | --- | 0.69 | 0.80 | 0.82 (0.70 ; 0.96) | 1.20E-02 | 0.81 | 0.75 (0.60 ; 0.93) | 7.94E-03 | 0.81 | 0.85 (0.70 ; 1.03) | 9.18E-02 |
| 2 | 237556343 | rs2600742 | C | G | 0.81 | 0.81 (0.73 ; 0.9) | 7.88E-05 | --- | 0.68 | 0.80 | 0.82 (0.70 ; 0.96) | 1.21E-02 | 0.81 | 0.75 (0.60 ; 0.92) | 7.14E-03 | 0.81 | 0.85 (0.70 ; 1.03) | 9.18E-02 |
| 2 | 237556353 | rs2600741 | A | G | 0.19 | 1.24 (1.11 ; 1.38) | 7.85E-05 | +++ | 0.68 | 0.20 | 1.22 (1.04 ; 1.43) | 1.21E-02 | 0.19 | 1.34 (1.08 ; 1.66) | 7.12E-03 | 0.19 | 1.18 (0.97 ; 1.43) | 9.18E-02 |
| 2 | 237557603 | rs2600739 | T | C | 0.67 | 0.78 (0.71 ; 0.85) | 7.79E-08 | --- | 0.81 | 0.67 | 0.76 (0.66 ; 0.87) | 4.41E-05 | 0.67 | 0.82 (0.68 ; 0.98) | 2.83E-02 | 0.68 | 0.79 (0.67 ; 0.93) | 5.25E-03 |
| 2 | 237560697 | rs2720115 | A | T | 0.67 | 0.79 (0.72 ; 0.86) | 2.23E-07 | --- | 0.84 | 0.67 | 0.77 (0.68 ; 0.88) | 1.48E-04 | 0.66 | 0.83 (0.69 ; 0.99) | 3.92E-02 | 0.67 | 0.78 (0.66 ; 0.92) | 3.52E-03 |
| 2 | 237561751 | rs2600737 | A | C | 0.67 | 0.79 (0.72 ; 0.86) | 1.51E-07 | --- | 0.87 | 0.67 | 0.77 (0.68 ; 0.88) | 1.45E-04 | 0.66 | 0.82 (0.68 ; 0.98) | 3.21E-02 | 0.67 | 0.78 (0.66 ; 0.92) | 2.98E-03 |
| 2 | 237562379 | rs1452052 | T | C | 0.33 | 1.27 (1.16 ; 1.39) | 2.31E-07 | +++ | 0.84 | 0.33 | 1.29 (1.13 ; 1.47) | 1.48E-04 | 0.34 | 1.21 (1.01 ; 1.45) | 3.97E-02 | 0.33 | 1.28 (1.08 ; 1.51) | 3.61E-03 |
| 2 | 237563161 | rs883362 | T | C | 0.67 | 0.79 (0.72 ; 0.86) | 1.25E-07 | --- | 0.81 | 0.67 | 0.77 (0.67 ; 0.88) | 8.21E-05 | 0.66 | 0.83 (0.69 ; 0.99) | 3.77E-02 | 0.67 | 0.78 (0.66 ; 0.92) | 3.48E-03 |

| | | | | | | | | | | | | | | | | | | |
|---|---|---|---|---|---|---|---|---|---|---|---|---|---|---|---|---|---|---|
| 2 | 237563328 | rs883363 | T | C | 0.67 | 0.79 (0.72 ; 0.86) | 1.54E-07 | --- | 0.82 | 0.67 | 0.77 (0.67 ; 0.88) | 1.01E-04 | 0.66 | 0.83 (0.69 ; 0.99) | 3.85E-02 | 0.67 | 0.78 (0.66 ; 0.92) | 3.48E-03 |
| 2 | 237563632 | rs2720117 | A | G | 0.67 | 0.79 (0.72 ; 0.86) | 2.54E-07 | --- | 0.79 | 0.67 | 0.77 (0.68 ; 0.88) | 1.14E-04 | 0.66 | 0.83 (0.70 ; 1.0) | 4.78E-02 | 0.67 | 0.78 (0.66 ; 0.93) | 4.11E-03 |
| 2 | 237563645 | rs2720118 | A | G | 0.66 | 0.79 (0.72 ; 0.86) | 1.76E-07 | --- | 0.83 | 0.66 | 0.77 (0.67 ; 0.88) | 9.80E-05 | 0.66 | 0.83 (0.69 ; 0.99) | 3.80E-02 | 0.67 | 0.79 (0.67 ; 0.93) | 4.23E-03 |
| 2 | 237564338 | rs2600736 | A | T | 0.33 | 1.27 (1.16 ; 1.39) | 2.31E-07 | +++ | 0.79 | 0.33 | 1.30 (1.14 ; 1.48) | 1.04E-04 | 0.34 | 1.20 (1.00 ; 1.44) | 4.74E-02 | 0.33 | 1.27 (1.08 ; 1.50) | 4.11E-03 |
| 2 | 237564716 | rs2720119 | C | G | 0.67 | 0.79 (0.72 ; 0.86) | 2.63E-07 | --- | 0.77 | 0.67 | 0.77 (0.68 ; 0.88) | 1.07E-04 | 0.66 | 0.84 (0.70 ; 1.0) | 5.18E-02 | 0.67 | 0.78 (0.66 ; 0.93) | 4.11E-03 |
| 2 | 237565290 | rs2720120 | A | C | 0.33 | 1.26 (1.15 ; 1.38) | 3.54E-07 | +++ | 0.79 | 0.33 | 1.29 (1.13 ; 1.47) | 1.71E-04 | 0.34 | 1.20 (1.00 ; 1.43) | 5.31E-02 | 0.33 | 1.28 (1.08 ; 1.51) | 3.50E-03 |
| 2 | 237565814 | rs2720121 | C | G | 0.33 | 1.26 (1.16 ; 1.38) | 3.20E-07 | +++ | 0.79 | 0.33 | 1.29 (1.13 ; 1.47) | 1.82E-04 | 0.34 | 1.20 (1.00 ; 1.43) | 5.16E-02 | 0.33 | 1.29 (1.09 ; 1.52) | 3.01E-03 |
| 2 | 237565901 | rs2600734 | T | C | 0.32 | 1.27 (1.16 ; 1.39) | 2.19E-07 | +++ | 0.74 | 0.32 | 1.32 (1.15 ; 1.51) | 5.10E-05 | 0.32 | 1.22 (1.01 ; 1.46) | 3.47E-02 | 0.31 | 1.24 (1.05 ; 1.47) | 1.02E-02 |
| 2 | 237565931 | rs2720122 | T | G | 0.33 | 1.27 (1.16 ; 1.39) | 1.73E-07 | +++ | 0.83 | 0.33 | 1.29 (1.13 ; 1.47) | 1.52E-04 | 0.34 | 1.21 (1.01 ; 1.45) | 3.92E-02 | 0.33 | 1.29 (1.09 ; 1.52) | 2.57E-03 |
| 2 | 237566315 | rs2600732 | A | C | 0.66 | 0.83 (0.76 ; 0.91) | 3.69E-05 | --- | 0.87 | 0.66 | 0.83 (0.73 ; 0.95) | 5.21E-03 | 0.65 | 0.86 (0.72 ; 1.03) | 9.40E-02 | 0.65 | 0.81 (0.68 ; 0.95) | 9.72E-03 |
| 2 | 237567023 | rs2720124 | T | G | 0.61 | 0.83 (0.76 ; 0.90) | 2.38E-05 | --- | 0.59 | 0.62 | 0.81 (0.71 ; 0.92) | 9.95E-04 | 0.61 | 0.90 (0.75 ; 1.07) | 2.24E-01 | 0.62 | 0.81 (0.69 ; 0.95) | 1.02E-02 |
| 2 | 237567309 | rs2720125 | T | G | 0.38 | 1.20 (1.10 ; 1.31) | 3.78E-05 | +++ | 0.45 | 0.38 | 1.24 (1.09 ; 1.41) | 1.11E-03 | 0.39 | 1.09 (0.91 ; 1.3) | 3.36E-01 | 0.38 | 1.24 (1.06 ; 1.46) | 8.15E-03 |
| 2 | 237567361 | rs2600731 | T | G | 0.62 | 0.83 (0.76 ; 0.91) | 4.14E-05 | --- | 0.46 | 0.62 | 0.81 (0.71 ; 0.92) | 1.14E-03 | 0.61 | 0.92 (0.77 ; 1.09) | 3.33E-01 | 0.62 | 0.81 (0.68 ; 0.95) | 9.02E-03 |
| 2 | 237567450 | rs2720126 | A | T | 0.38 | 1.20 (1.10 ; 1.31) | 4.43E-05 | +++ | 0.47 | 0.38 | 1.24 (1.09 ; 1.41) | 1.12E-03 | 0.39 | 1.09 (0.92 ; 1.3) | 3.31E-01 | 0.38 | 1.24 (1.05 ; 1.45) | 1.02E-02 |
| 2 | 237567494 | rs2720127 | A | G | 0.62 | 0.83 (0.77 ; 0.91) | 4.66E-05 | --- | 0.48 | 0.62 | 0.81 (0.71 ; 0.92) | 1.09E-03 | 0.61 | 0.92 (0.77 ; 1.09) | 3.30E-01 | 0.62 | 0.81 (0.69 ; 0.95) | 1.11E-02 |
| 2 | 237568329 | rs2600729 | T | C | 0.62 | 0.83 (0.76 ; 0.91) | 2.74E-05 | --- | 0.48 | 0.62 | 0.81 (0.71 ; 0.92) | 1.00E-03 | 0.61 | 0.91 (0.77 ; 1.09) | 2.99E-01 | 0.62 | 0.80 (0.68 ; 0.94) | 7.47E-03 |
| 2 | 237568342 | rs2720128 | T | C | 0.62 | 0.83 (0.76 ; 0.91) | 2.76E-05 | --- | 0.48 | 0.62 | 0.81 (0.71 ; 0.92) | 1.00E-03 | 0.61 | 0.91 (0.77 ; 1.09) | 3.01E-01 | 0.62 | 0.80 (0.68 ; 0.94) | 7.47E-03 |
| 2 | 237568361 | rs2600728 | A | T | 0.62 | 0.83 (0.76 ; 0.91) | 2.74E-05 | --- | 0.48 | 0.62 | 0.81 (0.71 ; 0.92) | 9.95E-04 | 0.61 | 0.91 (0.77 ; 1.09) | 3.00E-01 | 0.62 | 0.80 (0.68 ; 0.94) | 7.47E-03 |
| 2 | 237568487 | rs2600727 | A | C | 0.64 | 0.83 (0.76 ; 0.91) | 3.15E-05 | --- | 0.45 | 0.63 | 0.79 (0.70 ; 0.90) | 4.10E-04 | 0.63 | 0.91 (0.77 ; 1.09) | 2.99E-01 | 0.64 | 0.83 (0.70 ; 0.97) | 2.04E-02 |
| 2 | 237569002 | rs2720129 | T | G | 0.38 | 1.21 (1.11 ; 1.32) | 2.35E-05 | +++ | 0.46 | 0.38 | 1.24 (1.09 ; 1.41) | 8.92E-04 | 0.39 | 1.10 (0.92 ; 1.3) | 3.03E-01 | 0.38 | 1.25 (1.06 ; 1.47) | 6.78E-03 |
| 2 | 237569590 | rs921529 | A | T | 0.62 | 0.83 (0.76 ; 0.91) | 2.63E-05 | --- | 0.46 | 0.62 | 0.81 (0.71 ; 0.92) | 9.11E-04 | 0.61 | 0.91 (0.77 ; 1.09) | 3.06E-01 | 0.62 | 0.80 (0.68 ; 0.94) | 7.47E-03 |
| 2 | 237569810 | rs921528 | T | C | 0.62 | 0.83 (0.76 ; 0.91) | 2.59E-05 | --- | 0.47 | 0.62 | 0.81 (0.71 ; 0.92) | 9.04E-04 | 0.61 | 0.91 (0.77 ; 1.09) | 3.04E-01 | 0.62 | 0.80 (0.68 ; 0.94) | 7.47E-03 |
| 2 | 237570078 | rs2720130 | T | C | 0.62 | 0.83 (0.76 ; 0.91) | 2.59E-05 | --- | 0.47 | 0.62 | 0.81 (0.71 ; 0.92) | 9.06E-04 | 0.61 | 0.91 (0.77 ; 1.09) | 3.04E-01 | 0.62 | 0.80 (0.68 ; 0.94) | 7.47E-03 |
| 2 | 237570680 | rs2600724 | T | C | 0.36 | 1.21 (1.10 ; 1.32) | 2.91E-05 | +++ | 0.43 | 0.37 | 1.26 (1.11 ; 1.44) | 4.00E-04 | 0.37 | 1.10 (0.92 ; 1.31) | 3.06E-01 | 0.36 | 1.22 (1.03 ; 1.43) | 1.83E-02 |
| 2 | 237570723 | rs2720131 | A | G | 0.36 | 1.20 (1.10 ; 1.31) | 3.21E-05 | +++ | 0.44 | 0.37 | 1.26 (1.11 ; 1.44) | 4.01E-04 | 0.37 | 1.10 (0.92 ; 1.31) | 3.06E-01 | 0.36 | 1.21 (1.03 ; 1.42) | 2.04E-02 |
| 2 | 237571622 | rs2720132 | A | G | 0.62 | 0.83 (0.76 ; 0.91) | 3.51E-05 | --- | 0.42 | 0.62 | 0.80 (0.71 ; 0.91) | 8.37E-04 | 0.61 | 0.92 (0.77 ; 1.1) | 3.58E-01 | 0.62 | 0.81 (0.69 ; 0.95) | 8.80E-03 |
| 2 | 237571652 | rs2600723 | T | C | 0.36 | 1.20 (1.10 ; 1.31) | 3.54E-05 | +++ | 0.43 | 0.37 | 1.26 (1.11 ; 1.44) | 4.21E-04 | 0.37 | 1.09 (0.92 ; 1.3) | 3.18E-01 | 0.36 | 1.21 (1.03 ; 1.42) | 2.04E-02 |
| 2 | 237572620 | rs2720133 | T | C | 0.63 | 0.83 (0.76 ; 0.91) | 3.07E-05 | --- | 0.38 | 0.63 | 0.78 (0.69 ; 0.89) | 2.68E-04 | 0.63 | 0.92 (0.77 ; 1.09) | 3.26E-01 | 0.64 | 0.83 (0.71 ; 0.98) | 2.47E-02 |
| 2 | 237573038 | rs2720134 | A | G | 0.37 | 1.21 (1.10 ; 1.32) | 3.28E-05 | +++ | 0.38 | 0.37 | 1.27 (1.12 ; 1.45) | 2.90E-04 | 0.37 | 1.09 (0.92 ; 1.3) | 3.31E-01 | 0.36 | 1.21 (1.02 ; 1.42) | 2.41E-02 |
| 2 | 237573551 | rs921527 | A | G | 0.61 | 0.83 (0.76 ; 0.9) | 2.60E-05 | --- | 0.37 | 0.61 | 0.80 (0.70 ; 0.91) | 5.14E-04 | 0.61 | 0.93 (0.78 ; 1.1) | 3.87E-01 | 0.62 | 0.81 (0.69 ; 0.95) | 8.68E-03 |
| 8 | 111471061 | rs73314791 | A | G | 0.90 | 1.43 (1.23 ; 1.66) | 2.18E-06 | +++ | 0.75 | 0.89 | 1.50 (1.21 ; 1.86) | 2.31E-04 | 0.89 | 1.45 (1.08 ; 1.95) | 1.31E-02 | 0.91 | 1.30 (0.98 ; 1.74) | 6.95E-02 |
| 8 | 111481733 | rs16880731 | A | T | 0.90 | 1.44 (1.24 ; 1.67) | 1.60E-06 | +++ | 0.72 | 0.89 | 1.51 (1.22 ; 1.87) | 1.88E-04 | 0.90 | 1.46 (1.09 ; 1.97) | 1.12E-02 | 0.91 | 1.30 (0.98 ; 1.74) | 6.95E-02 |
| 8 | 111481995 | rs16880734 | A | G | 0.90 | 1.45 (1.25 ; 1.68) | 1.24E-06 | +++ | 0.74 | 0.89 | 1.51 (1.22 ; 1.88) | 1.71E-04 | 0.90 | 1.47 (1.09 ; 1.97) | 1.07E-02 | 0.91 | 1.31 (0.99 ; 1.75) | 6.25E-02 |
| 8 | 111484516 | rs13261067 | A | G | 0.90 | 1.45 (1.25 ; 1.68) | 1.23E-06 | +++ | 0.70 | 0.89 | 1.52 (1.23 ; 1.89) | 1.39E-04 | 0.90 | 1.46 (1.09 ; 1.97) | 1.12E-02 | 0.91 | 1.30 (0.98 ; 1.74) | 6.95E-02 |
| 8 | 111495105 | rs35314202 | A | T | 0.90 | 1.47 (1.26 ; 1.70) | 5.06E-07 | +++ | 0.71 | 0.89 | 1.54 (1.24 ; 1.91) | 1.00E-04 | 0.90 | 1.50 (1.11 ; 2.01) | 7.55E-03 | 0.91 | 1.32 (0.99 ; 1.76) | 5.56E-02 |
| 8 | 111501034 | rs13270632 | A | G | 0.90 | 1.48 (1.28 ; 1.72) | 2.72E-07 | +++ | 0.64 | 0.89 | 1.57 (1.26 ; 1.96) | 4.89E-05 | 0.90 | 1.50 (1.12 ; 2.02) | 7.28E-03 | 0.91 | 1.32 (0.99 ; 1.77) | 5.69E-02 |
| 8 | 111513400 | rs11991945 | A | G | 0.10 | 0.64 (0.55 ; 0.75) | 1.24E-08 | --- | 0.65 | 0.11 | 0.60 (0.48 ; 0.75) | 5.96E-06 | 0.10 | 0.66 (0.49 ; 0.89) | 6.59E-03 | 0.09 | 0.71 (0.53 ; 0.95) | 2.01E-02 |
| 8 | 111518434 | rs13265247 | A | G | 0.90 | 1.66 (1.42 ; 1.93) | 1.45E-10 | +++ | 0.39 | 0.89 | 1.83 (1.46 ; 2.30) | 1.89E-07 | 0.90 | 1.65 (1.21 ; 2.23) | 1.30E-03 | 0.91 | 1.41 (1.06 ; 1.89) | 1.91E-02 |
| 8 | 111521907 | rs71526761 | A | G | 0.91 | 1.73 (1.46 ; 2.05) | 2.13E-10 | +++ | 0.14 | 0.91 | 2.06 (1.60 ; 2.66) | 2.46E-08 | 0.91 | 1.65 (1.19 ; 2.28) | 2.77E-03 | 0.92 | 1.39 (1.01 ; 1.89) | 4.06E-02 |
| 8 | 111525870 | rs16880769 | A | C | 0.90 | 1.66 (1.42 ; 1.93) | 1.33E-10 | +++ | 0.39 | 0.89 | 1.83 (1.46 ; 2.30) | 1.82E-07 | 0.90 | 1.65 (1.22 ; 2.24) | 1.22E-03 | 0.91 | 1.41 (1.06 ; 1.89) | 1.90E-02 |
| 8 | 111530486 | rs67159506 | T | C | 0.10 | 0.60 (0.52 ; 0.70) | 1.36E-10 | --- | 0.38 | 0.11 | 0.55 (0.43 ; 0.69) | 1.87E-07 | 0.10 | 0.61 (0.45 ; 0.82) | 1.19E-03 | 0.09 | 0.71 (0.53 ; 0.95) | 1.95E-02 |
| 8 | 111533331 | rs73316466 | A | G | 0.89 | 1.63 (1.40 ; 1.90) | 2.49E-10 | +++ | 0.36 | 0.89 | 1.82 (1.46 ; 2.28) | 1.43E-07 | 0.89 | 1.58 (1.17 ; 2.12) | 2.77E-03 | 0.90 | 1.41 (1.06 ; 1.87) | 1.97E-02 |
| 8 | 111533799 | rs58445806 | A | C | 0.88 | 1.58 (1.37 ; 1.83) | 5.85E-10 | +++ | 0.51 | 0.88 | 1.70 (1.37 ; 2.10) | 1.02E-06 | 0.89 | 1.61 (1.21 ; 2.15) | 1.23E-03 | 0.89 | 1.39 (1.05 ; 1.83) | 2.03E-02 |
| 8 | 111535253 | rs35909617 | T | C | 0.89 | 1.60 (1.38 ; 1.85) | 3.78E-10 | +++ | 0.49 | 0.88 | 1.73 (1.39 ; 2.14) | 5.87E-07 | 0.89 | 1.60 (1.20 ; 2.15) | 1.45E-03 | 0.90 | 1.40 (1.06 ; 1.84) | 1.85E-02 |
| 8 | 111536392 | rs11997242 | A | G | 0.89 | 1.58 (1.37 ; 1.83) | 6.33E-10 | +++ | 0.55 | 0.88 | 1.69 (1.37 ; 2.10) | 1.30E-06 | 0.89 | 1.61 (1.21 ; 2.15) | 1.22E-03 | 0.90 | 1.39 (1.06 ; 1.84) | 1.86E-02 |
| 8 | 111536716 | rs11990209 | T | C | 0.16 | 0.75 (0.66 ; 0.84) | 2.45E-06 | --- | 0.63 | 0.17 | 0.72 (0.60 ; 0.86) | 2.69E-04 | 0.17 | 0.72 (0.56 ; 0.92) | 7.52E-03 | 0.15 | 0.82 (0.65 ; 1.04) | 9.89E-02 |
| 8 | 111538227 | rs11995877 | T | G | 0.89 | 1.58 (1.36 ; 1.83) | 8.41E-10 | +++ | 0.50 | 0.88 | 1.69 (1.37 ; 2.10) | 1.30E-06 | 0.89 | 1.61 (1.21 ; 2.15) | 1.22E-03 | 0.89 | 1.38 (1.04 ; 1.81) | 2.31E-02 |
| 8 | 111538735 | rs13255000 | C | G | 0.11 | 0.63 (0.55 ; 0.73) | 8.28E-10 | --- | 0.50 | 0.12 | 0.59 (0.48 ; 0.73) | 1.29E-06 | 0.11 | 0.62 (0.46 ; 0.83) | 1.22E-03 | 0.11 | 0.73 (0.55 ; 0.96) | 2.29E-02 |
| 8 | 111543667 | rs9942808 | T | C | 0.89 | 1.58 (1.37 ; 1.83) | 6.34E-10 | +++ | 0.55 | 0.88 | 1.69 (1.37 ; 2.09) | 1.29E-06 | 0.89 | 1.61 (1.21 ; 2.15) | 1.23E-03 | 0.90 | 1.39 (1.06 ; 1.84) | 1.86E-02 |
| 8 | 111544017 | rs11985542 | A | G | 0.89 | 1.58 (1.37 ; 1.83) | 6.33E-10 | +++ | 0.55 | 0.88 | 1.69 (1.37 ; 2.10) | 1.29E-06 | 0.89 | 1.61 (1.21 ; 2.15) | 1.23E-03 | 0.90 | 1.39 (1.06 ; 1.84) | 1.86E-02 |
| 8 | 111546388 | rs7002413 | A | T | 0.12 | 0.65 (0.57 ; 0.75) | 2.08E-09 | --- | 0.69 | 0.13 | 0.62 (0.50 ; 0.76) | 4.94E-06 | 0.12 | 0.65 (0.49 ; 0.85) | 1.98E-03 | 0.11 | 0.71 (0.55 ; 0.93) | 1.28E-02 |

| | | | | | | | | | | | | | | | | | | |
|---|---|---|---|---|---|---|---|---|---|---|---|---|---|---|---|---|---|---|
| 8 | 111547813 | rs16880800 | A | G | 0.89 | 1.59 (1.37 ; 1.84) | 6.47E-10 | +++ | 0.67 | 0.88 | 1.67 (1.35 ; 2.07) | 2.34E-06 | 0.89 | 1.61 (1.21 ; 2.15) | 1.27E-03 | 0.90 | 1.43 (1.08 ; 1.89) | 1.22E-02 |
| 8 | 111549604 | rs16880802 | A | T | 0.89 | 1.59 (1.38 ; 1.85) | 5.04E-10 | +++ | 0.80 | 0.88 | 1.66 (1.34 ; 2.06) | 3.37E-06 | 0.89 | 1.60 (1.20 ; 2.14) | 1.44E-03 | 0.90 | 1.48 (1.11 ; 1.96) | 6.72E-03 |
| 8 | 111553679 | rs16880808 | A | G | 0.89 | 1.56 (1.35 ; 1.81) | 1.88E-09 | +++ | 0.90 | 0.88 | 1.61 (1.30 ; 1.99) | 1.01E-05 | 0.89 | 1.56 (1.17 ; 2.09) | 2.27E-03 | 0.90 | 1.48 (1.12 ; 1.96) | 6.15E-03 |
| 8 | 111554872 | rs13280801 | A | G | 0.11 | 0.64 (0.55 ; 0.74) | 1.60E-09 | --- | 0.87 | 0.12 | 0.62 (0.50 ; 0.76) | 7.87E-06 | 0.11 | 0.64 (0.48 ; 0.85) | 2.20E-03 | 0.10 | 0.68 (0.51 ; 0.90) | 6.72E-03 |
| 8 | 111557094 | rs726685 | A | G | 0.89 | 1.57 (1.35 ; 1.81) | 1.65E-09 | +++ | 0.86 | 0.88 | 1.62 (1.31 ; 2.01) | 7.89E-06 | 0.89 | 1.57 (1.18 ; 2.09) | 2.20E-03 | 0.90 | 1.47 (1.11 ; 1.95) | 6.91E-03 |
| 8 | 111558924 | rs13250442 | A | G | 0.11 | 0.64 (0.55 ; 0.74) | 1.59E-09 | --- | 0.87 | 0.12 | 0.62 (0.50 ; 0.76) | 7.93E-06 | 0.11 | 0.64 (0.48 ; 0.85) | 2.16E-03 | 0.10 | 0.68 (0.51 ; 0.90) | 6.72E-03 |
| 8 | 111559103 | rs13251031 | T | G | 0.11 | 0.63 (0.55 ; 0.73) | 1.22E-09 | --- | 0.88 | 0.12 | 0.61 (0.50 ; 0.76) | 6.90E-06 | 0.11 | 0.64 (0.48 ; 0.85) | 2.16E-03 | 0.10 | 0.67 (0.51 ; 0.89) | 5.93E-03 |
| 8 | 111562304 | rs13265535 | T | C | 0.89 | 1.57 (1.35 ; 1.82) | 1.65E-09 | +++ | 0.86 | 0.88 | 1.62 (1.31 ; 2.01) | 7.90E-06 | 0.89 | 1.57 (1.18 ; 2.09) | 2.15E-03 | 0.90 | 1.47 (1.11 ; 1.95) | 7.01E-03 |
| 8 | 111563316 | rs11994178 | A | G | 0.89 | 1.56 (1.35 ; 1.81) | 1.98E-09 | +++ | 0.91 | 0.88 | 1.61 (1.30 ; 1.99) | 1.16E-05 | 0.89 | 1.57 (1.18 ; 2.09) | 2.20E-03 | 0.90 | 1.49 (1.12 ; 1.97) | 5.88E-03 |
| 8 | 111563468 | rs13273764 | A | T | 0.89 | 1.56 (1.35 ; 1.81) | 1.98E-09 | +++ | 0.91 | 0.88 | 1.61 (1.30 ; 1.99) | 1.16E-05 | 0.89 | 1.57 (1.18 ; 2.09) | 2.20E-03 | 0.90 | 1.49 (1.12 ; 1.97) | 5.88E-03 |
| 8 | 111567651 | rs13278891 | A | G | 0.89 | 1.57 (1.35 ; 1.82) | 1.63E-09 | +++ | 0.87 | 0.88 | 1.62 (1.31 ; 2.01) | 7.90E-06 | 0.89 | 1.57 (1.17 ; 2.09) | 2.23E-03 | 0.90 | 1.48 (1.11 ; 1.96) | 6.72E-03 |
| 8 | 111567783 | rs13279857 | A | G | 0.11 | 0.64 (0.55 ; 0.74) | 1.46E-09 | --- | 0.86 | 0.12 | 0.61 (0.50 ; 0.76) | 6.91E-06 | 0.11 | 0.64 (0.48 ; 0.85) | 2.27E-03 | 0.10 | 0.68 (0.51 ; 0.90) | 6.72E-03 |
| 8 | 111568197 | rs13252314 | A | G | 0.89 | 1.57 (1.35 ; 1.82) | 1.63E-09 | +++ | 0.87 | 0.88 | 1.62 (1.31 ; 2.01) | 7.90E-06 | 0.89 | 1.57 (1.17 ; 2.09) | 2.23E-03 | 0.90 | 1.48 (1.11 ; 1.96) | 6.72E-03 |
| 8 | 111568286 | rs13253273 | A | G | 0.11 | 0.64 (0.55 ; 0.74) | 1.63E-09 | --- | 0.87 | 0.12 | 0.62 (0.50 ; 0.76) | 7.90E-06 | 0.11 | 0.64 (0.48 ; 0.85) | 2.23E-03 | 0.10 | 0.68 (0.51 ; 0.90) | 6.72E-03 |
| 8 | 111572193 | rs7828750 | A | G | 0.11 | 0.64 (0.55 ; 0.74) | 1.63E-09 | --- | 0.87 | 0.12 | 0.62 (0.50 ; 0.76) | 7.92E-06 | 0.11 | 0.64 (0.48 ; 0.85) | 2.23E-03 | 0.10 | 0.68 (0.51 ; 0.90) | 6.72E-03 |
| 8 | 111575206 | rs1494243 | A | G | 0.89 | 1.56 (1.35 ; 1.81) | 1.83E-09 | +++ | 0.90 | 0.88 | 1.61 (1.30 ; 1.99) | 1.01E-05 | 0.89 | 1.56 (1.17 ; 2.08) | 2.31E-03 | 0.90 | 1.49 (1.12 ; 1.97) | 5.88E-03 |
| 8 | 111576073 | rs73320482 | A | G | 0.89 | 1.57 (1.35 ; 1.81) | 1.64E-09 | +++ | 0.87 | 0.88 | 1.62 (1.31 ; 2.01) | 7.94E-06 | 0.89 | 1.57 (1.17 ; 2.09) | 2.23E-03 | 0.90 | 1.48 (1.11 ; 1.96) | 6.72E-03 |
| 8 | 111576851 | rs67856195 | A | G | 0.89 | 1.57 (1.35 ; 1.81) | 1.64E-09 | +++ | 0.87 | 0.88 | 1.62 (1.31 ; 2.01) | 7.93E-06 | 0.89 | 1.57 (1.17 ; 2.09) | 2.23E-03 | 0.90 | 1.48 (1.11 ; 1.96) | 6.72E-03 |
| 8 | 111578094 | rs34642482 | A | G | 0.89 | 1.57 (1.36 ; 1.82) | 1.55E-09 | +++ | 0.87 | 0.88 | 1.62 (1.31 ; 2.01) | 7.94E-06 | 0.89 | 1.57 (1.17 ; 2.09) | 2.23E-03 | 0.90 | 1.48 (1.12 ; 1.96) | 6.37E-03 |
| 8 | 111578478 | rs35697720 | A | G | 0.89 | 1.56 (1.35 ; 1.81) | 1.91E-09 | +++ | 0.90 | 0.88 | 1.61 (1.30 ; 1.99) | 1.01E-05 | 0.89 | 1.56 (1.17 ; 2.08) | 2.35E-03 | 0.90 | 1.48 (1.12 ; 1.97) | 6.06E-03 |
| 8 | 111579239 | rs11997872 | T | C | 0.11 | 0.64 (0.55 ; 0.74) | 1.64E-09 | --- | 0.87 | 0.12 | 0.62 (0.50 ; 0.76) | 7.94E-06 | 0.11 | 0.64 (0.48 ; 0.85) | 2.23E-03 | 0.10 | 0.68 (0.51 ; 0.90) | 6.72E-03 |
| 8 | 111580170 | rs1494232 | T | G | 0.11 | 0.64 (0.55 ; 0.74) | 1.83E-09 | --- | 0.90 | 0.12 | 0.62 (0.50 ; 0.77) | 1.01E-05 | 0.11 | 0.64 (0.48 ; 0.85) | 2.31E-03 | 0.10 | 0.67 (0.51 ; 0.89) | 5.88E-03 |
| 8 | 111581847 | rs1389671 | T | G | 0.11 | 0.64 (0.55 ; 0.74) | 1.63E-09 | --- | 0.87 | 0.12 | 0.62 (0.50 ; 0.76) | 7.92E-06 | 0.11 | 0.64 (0.48 ; 0.85) | 2.23E-03 | 0.10 | 0.68 (0.51 ; 0.90) | 6.72E-03 |
| 8 | 111584146 | rs36110122 | T | C | 0.11 | 0.64 (0.55 ; 0.74) | 2.09E-09 | --- | 0.91 | 0.12 | 0.62 (0.50 ; 0.77) | 1.16E-05 | 0.11 | 0.64 (0.48 ; 0.85) | 2.32E-03 | 0.10 | 0.67 (0.51 ; 0.89) | 5.88E-03 |
| 8 | 111586155 | rs13277548 | T | G | 0.11 | 0.64 (0.55 ; 0.74) | 2.06E-09 | --- | 0.91 | 0.12 | 0.62 (0.50 ; 0.77) | 1.16E-05 | 0.11 | 0.64 (0.48 ; 0.85) | 2.29E-03 | 0.10 | 0.67 (0.51 ; 0.89) | 5.88E-03 |
| 8 | 111586510 | rs58672526 | A | G | 0.11 | 0.64 (0.55 ; 0.74) | 2.06E-09 | --- | 0.91 | 0.12 | 0.62 (0.50 ; 0.77) | 1.16E-05 | 0.11 | 0.64 (0.48 ; 0.85) | 2.29E-03 | 0.10 | 0.67 (0.51 ; 0.89) | 5.88E-03 |
| 8 | 111587121 | rs13252162 | A | G | 0.11 | 0.64 (0.55 ; 0.74) | 1.67E-09 | --- | 0.87 | 0.12 | 0.62 (0.50 ; 0.76) | 7.96E-06 | 0.11 | 0.64 (0.48 ; 0.85) | 2.28E-03 | 0.10 | 0.68 (0.51 ; 0.90) | 6.72E-03 |
| 8 | 111595769 | rs4486175 | A | G | 0.89 | 1.51 (1.31 ; 1.75) | 1.94E-08 | +++ | 0.96 | 0.88 | 1.51 (1.23 ; 1.86) | 1.02E-04 | 0.89 | 1.56 (1.17 ; 2.08) | 2.44E-03 | 0.90 | 1.48 (1.11 ; 1.96) | 6.72E-03 |
| 8 | 111598824 | rs16880887 | A | G | 0.11 | 0.66 (0.57 ; 0.76) | 1.97E-08 | --- | 0.96 | 0.12 | 0.66 (0.54 ; 0.82) | 1.03E-04 | 0.11 | 0.64 (0.48 ; 0.86) | 2.48E-03 | 0.10 | 0.68 (0.51 ; 0.90) | 6.72E-03 |
| 8 | 111600446 | rs34880112 | T | C | 0.12 | 0.66 (0.57 ; 0.77) | 2.29E-08 | --- | 0.97 | 0.12 | 0.67 (0.54 ; 0.82) | 1.43E-04 | 0.11 | 0.64 (0.48 ; 0.86) | 2.49E-03 | 0.11 | 0.67 (0.51 ; 0.89) | 5.55E-03 |
| 8 | 111603503 | rs75741277 | T | G | 0.11 | 0.66 (0.57 ; 0.76) | 2.13E-08 | --- | 0.96 | 0.12 | 0.66 (0.54 ; 0.82) | 1.03E-04 | 0.11 | 0.64 (0.48 ; 0.85) | 2.44E-03 | 0.10 | 0.68 (0.51 ; 0.90) | 7.33E-03 |
| 8 | 111609469 | rs11992948 | A | G | 0.12 | 0.68 (0.59 ; 0.78) | 8.91E-08 | --- | 0.95 | 0.12 | 0.66 (0.54 ; 0.82) | 1.02E-04 | 0.12 | 0.70 (0.53 ; 0.92) | 1.21E-02 | 0.10 | 0.68 (0.51 ; 0.90) | 6.91E-03 |
| 8 | 111611907 | rs7016455 | A | T | 0.12 | 0.68 (0.59 ; 0.78) | 8.75E-08 | --- | 0.95 | 0.12 | 0.66 (0.54 ; 0.82) | 1.03E-04 | 0.11 | 0.70 (0.53 ; 0.92) | 1.21E-02 | 0.10 | 0.68 (0.51 ; 0.90) | 6.72E-03 |
| 8 | 111612360 | rs34077261 | T | C | 0.09 | 0.64 (0.54 ; 0.75) | 1.60E-07 | --- | 0.70 | 0.09 | 0.59 (0.46 ; 0.76) | 3.31E-05 | 0.09 | 0.68 (0.49 ; 0.94) | 1.78E-02 | 0.08 | 0.68 (0.50 ; 0.94) | 2.09E-02 |
| 8 | 111613586 | rs6984924 | T | C | 0.88 | 1.48 (1.28 ; 1.71) | 8.86E-08 | +++ | 0.95 | 0.88 | 1.51 (1.23 ; 1.86) | 1.03E-04 | 0.89 | 1.43 (1.08 ; 1.89) | 1.19E-02 | 0.90 | 1.47 (1.11 ; 1.95) | 6.91E-03 |
| 8 | 111614642 | rs66906875 | T | C | 0.88 | 1.49 (1.29 ; 1.71) | 6.80E-08 | +++ | 0.95 | 0.88 | 1.51 (1.23 ; 1.86) | 1.03E-04 | 0.89 | 1.43 (1.08 ; 1.89) | 1.19E-02 | 0.90 | 1.50 (1.13 ; 1.99) | 5.18E-03 |
| 8 | 111615297 | rs6995161 | T | G | 0.89 | 1.49 (1.29 ; 1.72) | 6.64E-08 | +++ | 0.95 | 0.88 | 1.51 (1.23 ; 1.86) | 1.03E-04 | 0.89 | 1.43 (1.08 ; 1.89) | 1.19E-02 | 0.90 | 1.50 (1.13 ; 1.99) | 5.03E-03 |
| 8 | 111615743 | rs6469316 | T | C | 0.89 | 1.49 (1.29 ; 1.71) | 6.77E-08 | +++ | 0.95 | 0.88 | 1.51 (1.23 ; 1.86) | 1.04E-04 | 0.89 | 1.43 (1.08 ; 1.89) | 1.21E-02 | 0.90 | 1.50 (1.13 ; 1.99) | 5.03E-03 |
| 8 | 111616532 | rs6980765 | A | G | 0.88 | 1.48 (1.29 ; 1.71) | 7.07E-08 | +++ | 0.95 | 0.88 | 1.51 (1.23 ; 1.86) | 1.04E-04 | 0.89 | 1.43 (1.08 ; 1.89) | 1.20E-02 | 0.90 | 1.49 (1.13 ; 1.98) | 5.35E-03 |
| 8 | 111618037 | rs34517324 | A | T | 0.14 | 0.78 (0.68 ; 0.88) | 9.29E-05 | --- | 0.37 | 0.15 | 0.70 (0.58 ; 0.85) | 2.88E-04 | 0.14 | 0.82 (0.64 ; 1.05) | 1.09E-01 | 0.14 | 0.86 (0.68 ; 1.09) | 2.15E-01 |
| 8 | 111620778 | rs11991599 | A | G | 0.86 | 1.29 (1.14 ; 1.46) | 8.83E-05 | +++ | 0.38 | 0.85 | 1.42 (1.17 ; 1.72) | 3.22E-04 | 0.86 | 1.24 (0.97 ; 1.59) | 8.76E-02 | 0.86 | 1.15 (0.91 ; 1.46) | 2.32E-01 |
| 8 | 111624563 | rs4089010 | T | C | 0.86 | 1.29 (1.14 ; 1.47) | 8.26E-05 | +++ | 0.40 | 0.85 | 1.42 (1.17 ; 1.72) | 3.23E-04 | 0.86 | 1.24 (0.97 ; 1.59) | 8.75E-02 | 0.86 | 1.16 (0.92 ; 1.46) | 2.20E-01 |
| 8 | 111627180 | rs11985995 | A | C | 0.14 | 0.77 (0.68 ; 0.88) | 8.45E-05 | --- | 0.39 | 0.15 | 0.71 (0.58 ; 0.85) | 3.23E-04 | 0.14 | 0.80 (0.63 ; 1.03) | 8.76E-02 | 0.14 | 0.86 (0.68 ; 1.09) | 2.24E-01 |
| 8 | 111628745 | rs1353209 | A | G | 0.86 | 1.29 (1.14 ; 1.47) | 8.13E-05 | +++ | 0.40 | 0.85 | 1.42 (1.17 ; 1.72) | 3.20E-04 | 0.86 | 1.24 (0.97 ; 1.59) | 8.75E-02 | 0.86 | 1.16 (0.92 ; 1.46) | 2.19E-01 |
| 8 | 111631522 | rs11990174 | A | G | 0.14 | 0.77 (0.68 ; 0.87) | 5.55E-05 | --- | 0.37 | 0.15 | 0.70 (0.57 ; 0.84) | 1.97E-04 | 0.14 | 0.81 (0.63 ; 1.04) | 1.03E-01 | 0.13 | 0.85 (0.67 ; 1.08) | 1.91E-01 |
| 8 | 111633642 | rs7015828 | A | G | 0.14 | 0.77 (0.68 ; 0.87) | 5.59E-05 | --- | 0.37 | 0.15 | 0.70 (0.57 ; 0.84) | 1.99E-04 | 0.14 | 0.81 (0.63 ; 1.04) | 1.03E-01 | 0.13 | 0.85 (0.67 ; 1.08) | 1.91E-01 |
| 8 | 111635212 | rs6987732 | A | G | 0.14 | 0.78 (0.68 ; 0.88) | 9.85E-05 | --- | 0.46 | 0.15 | 0.71 (0.59 ; 0.86) | 4.30E-04 | 0.14 | 0.81 (0.63 ; 1.04) | 1.03E-01 | 0.13 | 0.86 (0.68 ; 1.08) | 1.95E-01 |
| 8 | 111636762 | rs16880926 | A | G | 0.86 | 1.29 (1.13 ; 1.46) | 9.53E-05 | +++ | 0.46 | 0.85 | 1.41 (1.16 ; 1.70) | 4.29E-04 | 0.86 | 1.23 (0.96 ; 1.59) | 1.03E-01 | 0.87 | 1.17 (0.92 ; 1.48) | 1.91E-01 |
| 8 | 111660606 | rs34379073 | A | G | 0.09 | 0.68 (0.57 ; 0.8) | 3.27E-06 | --- | 0.71 | 0.09 | 0.63 (0.49 ; 0.80) | 1.90E-04 | 0.09 | 0.74 (0.54 ; 1.01) | 6.03E-02 | 0.08 | 0.69 (0.50 ; 0.96) | 2.71E-02 |
| 8 | 111684296 | rs13254072 | A | C | 0.91 | 1.51 (1.28 ; 1.78) | 9.72E-07 | +++ | 0.78 | 0.91 | 1.60 (1.26 ; 2.04) | 1.26E-04 | 0.91 | 1.40 (1.02 ; 1.92) | 3.48E-02 | 0.92 | 1.46 (1.06 ; 2.02) | 2.12E-02 |
| 8 | 111691751 | rs35429403 | T | C | 0.08 | 0.70 (0.59 ; 0.83) | 3.10E-05 | --- | 0.55 | 0.09 | 0.63 (0.49 ; 0.81) | 3.50E-04 | 0.08 | 0.72 (0.52 ; 1.0) | 4.81E-02 | 0.07 | 0.80 (0.57 ; 1.10) | 1.70E-01 |

**Supplementary Table 9: eQTL lookup and colocalization at *ACKR3* locus.**

GTEx database (v10) was interrogated for significant eQTL for rs2708184, the lead SNP at *ACKR3* locus. All significant associations are indicated. For each association colocalizaiton between hEDS association and eQTL associaiton was tested using coloc R package. N SNPs indicates the number of SNPs shared between the two association studies in the locus (leadSNP +/- 500kb). PP.H0.abf to PP.H4.abf columns respectively indicate the computed probabilities for: no association at the locus (H0), association for trait 1 (hEDS) only at the locus (H1), association for trait 2 (eQTL) only at the locus (H2), associations for both traits at the locus but with different causal variants (H3), association of both [illegible]

| GENE | | | | | | TISSUE | Colocalization | | | | | |
|---|---|---|---|---|---|---|---|---|---|---|---|---|
| **ID** | **Name** | **Chr** | **start** | **end** | **strand** | **Tissue** | **N SNPs** | **PP.H0.abf** | **PP.H1.abf** | **PP.H2.abf** | **PP.H3.abf** | **PP.H4.abf** |
| ENSG00000144476 | ACKR3 | chr2 | 236567787 | 236582354 | + | Nerve_Tibial | 4050 | 0% | 0% | 0% | 14% | 86% |
| ENSG00000144476 | ACKR3 | chr2 | 236567787 | 236582354 | + | Cells_Cultured_fibroblasts | 4050 | 0% | 0% | 0% | 96% | 4% |
| ENSG00000144476 | ACKR3 | chr2 | 236567787 | 236582354 | + | Artery_Tibial | 4050 | 0% | 0% | 0% | 91% | 8% |
| ENSG00000144476 | ACKR3 | chr2 | 236567787 | 236582354 | + | Adipose_Subcutaneous | 4050 | 0% | 0% | 0% | 91% | 9% |

**Supplementary Table 10: Predictions of transcription factor binding sites affected by rs2600746 genotype**

Hocomoco core database of transcription factor binding motifs (v13) was interrogated for differential predicted similarity with 51 bp DNA region centered on rs2600746, using PERFECTOS-APE webserver, with default parameters. *P* -value of similarity with TF motif is indicated for hEDS risk allele (C) and protective allele (G).

| **Motif** | **P-value C allele** | **P-value G allele** | **Fold change** | **Effect of hEDS risk allele (C)** |
|---|---|---|---|---|
| AHR.H13CORE.0.P.B | 3.3E-05 | 2.4E-03 | 72 | UP |
| ZN335.H13CORE.1.P.D | 3.7E-03 | 6.1E-05 | 60 | DOWN |
| CXXC4.H13CORE.0.PSGIB.A | 1.0E-04 | 5.1E-03 | 51 | UP |
| Z780A.H13CORE.0.P.C | 2.0E-03 | 6.1E-05 | 33 | DOWN |
| MGAP.H13CORE.1.S.B | 4.8E-04 | 7.6E-03 | 16 | UP |
| ZN75D.H13CORE.0.P.B | 1.1E-04 | 1.7E-03 | 16 | UP |
| ZN611.H13CORE.0.P.C | 5.7E-03 | 3.8E-04 | 15 | DOWN |
| HIF3A.H13CORE.1.P.C | 3.4E-04 | 4.2E-03 | 12 | UP |
| RFX6.H13CORE.0.P.C | 4.9E-03 | 4.4E-04 | 11 | DOWN |
| ZSCA4.H13CORE.0.SM.B | 4.9E-04 | 5.2E-03 | 11 | UP |
| ZN33B.H13CORE.0.P.C | 4.3E-03 | 4.0E-04 | 11 | DOWN |
| ESR2.H13CORE.0.P.B | 4.8E-03 | 4.9E-04 | 10 | DOWN |
| HES1.H13CORE.1.S.C | 1.8E-03 | 2.5E-04 | 7 | DOWN |
| PRDM4.H13CORE.0.PSM.A | 5.6E-04 | 8.4E-05 | 7 | DOWN |
| ZN322.H13CORE.0.P.C | 1.0E-03 | 1.6E-04 | 6 | DOWN |
| ZN557.H13CORE.0.P.C | 6.9E-04 | 1.2E-04 | 6 | DOWN |
| AHRR.H13CORE.0.P.C | 4.0E-05 | 2.0E-04 | 5 | UP |
| HES1.H13CORE.0.S.C | 1.0E-03 | 2.1E-04 | 5 | DOWN |
| ZNF48.H13CORE.0.PSG.A | 5.0E-04 | 2.3E-03 | 5 | UP |
| ZBT14.H13CORE.0.P.C | 4.2E-04 | 1.9E-03 | 5 | UP |
| ZN880.H13CORE.0.P.C | 6.6E-04 | 1.5E-04 | 5 | DOWN |
| ZN180.H13CORE.0.P.C | 3.6E-04 | 1.7E-03 | 5 | UP |
| FOXG1.H13CORE.0.PSM.A | 1.1E-05 | 5.1E-05 | 4 | UP |
| MGA.H13CORE.0.PSG.A | 4.5E-04 | 2.0E-03 | 4 | UP |
| ZN570.H13CORE.0.P.C | 2.0E-03 | 4.7E-04 | 4 | DOWN |
| PAX7.H13CORE.0.P.B | 3.7E-04 | 1.5E-03 | 4 | UP |

**Supplementary Table 11. Top gene-based association results for hEDS (meta-analysis) using LDAK-GBAT.**

For each autosomal gene (RefSeq, hg19/GRCh37) we report: chromosome and coordinates; the number of SNPs within gene bounds before QC (N SNPs) and retained in the test after QC and reference overlap (N SNPs used); the minimum single-variant P-value observed within the gene (Gene_SNP_MinP) and its Bonferroni adjustment as reported by LDAK-GBAT (Gene_SNP_MinP_Bonferroni); the estimated gene-level SNP heritability and standard error (Gene_h2, Gene_h2_se); the LDAK-GBAT Z statistic (GBAT_ZSTAT) and empirical permutation P-value (GBAT_P); and multiple-testing adjusted P-values across all tested genes (Bonferroni over 17,441 genes; Benjamini–Hochberg FDR). "clump = yes" marks LD-independent index genes retained after LDAK-GBAT clumping (same-chromosome pairs with $r^2$>0.1 are grouped and only the most significant gene is labeled "yes"); blank entries are non-index members of a clumped set. Analyses used the Human Default Model with MAF weighting $[MAF\times(1–MAF)]^{0.75}$, a 10,000-sample UK Biobank reference panel (12.8M imputed [illegible]

| GENE | CHR | START | STOP | N SNPS | N SNPs used | Gene SNP MinP | Gene SNP MinP Bonferroni | Gene h2 | Gene h2 se | GBAT ZSTAT | GBAT P | clump | GBAT P Bonferroni | GBAT P FDR |
|---|---|---|---|---|---|---|---|---|---|---|---|---|---|---|
| PXDNL | 8 | 52232137 | 52722005 | 1225 | 105 | 6.6E-06 | 8.1E-03 | 0.055 | 0.041 | 5.28 | 6.3E-08 | yes | 0.001 | 0.001 |
| ACACA | 17 | 35441927 | 35766902 | 178 | 35 | 1.8E-04 | 3.2E-02 | 0.025 | 0.013 | 4.81 | 7.6E-07 | yes | 0.014 | 0.007 |
| SLC39A13 | 11 | 47428683 | 47438051 | 20 | 7 | 4.8E-06 | 9.7E-05 | 0.010 | 0.007 | 4.71 | 1.2E-06 | yes | 0.022 | 0.007 |
| NPFFR2 | 4 | 72897521 | 73013918 | 95 | 17 | 1.9E-05 | 1.8E-03 | 0.040 | 0.025 | 4.64 | 1.8E-06 | yes | 0.031 | 0.007 |
| PDGFRL | 8 | 17433942 | 17500642 | 365 | 55 | 7.5E-07 | 2.7E-04 | 0.060 | 0.048 | 4.62 | 2.0E-06 | yes | 0.035 | 0.007 |
| MADD | 11 | 47290927 | 47351582 | 118 | 20 | 9.0E-06 | 1.1E-03 | 0.007 | 0.004 | 4.41 | 5.3E-06 | | 0.094 | 0.016 |
| SPI1 | 11 | 47376409 | 47400127 | 56 | 13 | 1.4E-06 | 7.9E-05 | 0.006 | 0.004 | 4.29 | 9.1E-06 | | 0.161 | 0.023 |
| SNRPE | 1 | 2.04E+08 | 203840280 | 30 | 7 | 2.5E-06 | 7.6E-05 | 0.005 | 0.004 | 4.14 | 1.7E-05 | | 0.310 | 0.039 |
| ZC3H11A | 1 | 2.04E+08 | 203823256 | 130 | 11 | 5.4E-06 | 7.0E-04 | 0.016 | 0.027 | 4.11 | 2.0E-05 | | 0.352 | 0.039 |
| NR1H3 | 11 | 47269851 | 47290584 | 40 | 10 | 1.9E-05 | 7.6E-04 | 0.005 | 0.003 | 4.01 | 3.1E-05 | | 0.55 | 0.052 |
| NIFK | 2 | 1.22E+08 | 122494503 | 8 | 3 | 6.5E-05 | 5.2E-04 | 0.004 | 0.003 | 3.99 | 3.3E-05 | | 0.58 | 0.052 |
| MTCH2 | 11 | 47638858 | 47664206 | 17 | 9 | 5.6E-06 | 9.5E-05 | 0.005 | 0.004 | 3.92 | 4.4E-05 | | 0.78 | 0.052 |
| RPS10 | 6 | 34385231 | 34393902 | 12 | 5 | 2.0E-04 | 2.4E-03 | 0.003 | 0.003 | 3.91 | 4.5E-05 | | 0.81 | 0.052 |
| PLCE1 | 10 | 95753746 | 96088149 | 537 | 74 | 2.2E-05 | 1.2E-02 | 0.009 | 0.005 | 3.90 | 4.9E-05 | | 0.87 | 0.052 |
| GC | 4 | 72607410 | 72671237 | 125 | 17 | 1.4E-06 | 1.8E-04 | 0.006 | 0.004 | 3.88 | 5.3E-05 | | 0.93 | 0.052 |
| ZNF33B | 10 | 43084532 | 43134016 | 97 | 12 | 1.2E-04 | 1.2E-02 | 0.005 | 0.004 | 3.88 | 5.3E-05 | | 0.93 | 0.052 |
| IL16 | 15 | 81474941 | 81605104 | 306 | 31 | 8.2E-04 | 2.2E-01 | 0.014 | 0.008 | 3.86 | 5.6E-05 | | 0.99 | 0.052 |
| DEFB116 | 20 | 29891015 | 29896388 | 9 | 4 | 1.3E-04 | 1.1E-03 | 0.002 | 0.002 | 3.86 | 5.7E-05 | | 1.00 | 0.052 |
| PIK3AP1 | 10 | 98353069 | 98480279 | 233 | 43 | 3.6E-04 | 8.1E-02 | 0.012 | 0.007 | 3.85 | 5.9E-05 | | 1.00 | 0.052 |
| ZNF547 | 19 | 57874891 | 57890927 | 26 | 7 | 5.2E-04 | 1.3E-02 | 0.007 | 0.005 | 3.85 | 5.9E-05 | | 1.00 | 0.052 |
| CLEC18A | 16 | 69984608 | 69998250 | 1 | 1 | 3.1E-05 | 3.1E-05 | 0.003 | 0.004 | 3.83 | 6.5E-05 | | 1.00 | 0.054 |
| PSMC3 | 11 | 47440320 | 47448024 | 12 | 6 | 1.3E-03 | 1.6E-02 | 0.009 | 0.007 | 3.81 | 7.0E-05 | | 1.00 | 0.054 |
| ZNF511 | 10 | 1.35E+08 | 135126666 | 4 | 3 | 1.9E-04 | 7.5E-04 | 0.004 | 0.004 | 3.81 | 7.0E-05 | | 1.00 | 0.054 |
| ZNF304 | 19 | 57862645 | 57871266 | 21 | 11 | 3.0E-04 | 6.2E-03 | 0.012 | 0.009 | 3.77 | 8.3E-05 | | 1.00 | 0.06 |
| ACSL6 | 5 | 1.31E+08 | 131347761 | 63 | 13 | 5.5E-06 | 3.4E-04 | 0.009 | 0.007 | 3.75 | 8.9E-05 | | 1.00 | 0.06 |
| MYBPC3 | 11 | 47352957 | 47374253 | 38 | 16 | 4.3E-06 | 1.6E-04 | 0.005 | 0.003 | 3.72 | 1.0E-04 | | 1.00 | 0.07 |
| CELF1 | 11 | 47487489 | 47574792 | 79 | 24 | 5.2E-06 | 4.1E-04 | 0.005 | 0.003 | 3.71 | 1.0E-04 | | 1.00 | 0.07 |
| ZNF57 | 19 | 2900896 | 2918641 | 64 | 8 | 6.4E-04 | 4.0E-02 | 0.031 | 0.023 | 3.71 | 1.1E-04 | | 1.00 | 0.07 |
| DACT2 | 6 | 1.69E+08 | 168720459 | 73 | 25 | 2.0E-06 | 1.5E-04 | 0.006 | 0.003 | 3.70 | 1.1E-04 | | 1.00 | 0.07 |

| | | | | | | | | | | | | | | |
|---|---|---|---|---|---|---|---|---|---|---|---|---|---|---|
| CFDP1 | 16 | 75327608 | 75467387 | 373 | 23 | 1.1E-06 | 4.2E-04 | 0.003 | 0.002 | 3.68 | 1.2E-04 | | 1.00 | 0.07 |
| ESR1 | 6 | 1.52E+08 | 152424409 | 1037 | 120 | 1.5E-04 | 1.5E-01 | 0.006 | 0.003 | 3.66 | 1.3E-04 | | 1.00 | 0.07 |
| SIRT6 | 19 | 4174106 | 4182596 | 10 | 5 | 3.0E-05 | 3.0E-04 | 0.004 | 0.003 | 3.65 | 1.3E-04 | | 1.00 | 0.07 |
| KREMEN1 | 22 | 29469066 | 29564321 | 187 | 27 | 1.0E-05 | 1.9E-03 | 0.011 | 0.009 | 3.62 | 1.5E-04 | | 1.00 | 0.08 |
| TEDDM1 | 1 | 1.82E+08 | 182369751 | 7 | 3 | 8.7E-05 | 6.1E-04 | 0.003 | 0.003 | 3.62 | 1.5E-04 | | 1.00 | 0.08 |
| FOXE3 | 1 | 47881744 | 47883724 | 2 | 2 | 3.1E-05 | 6.2E-05 | 0.004 | 0.005 | 3.57 | 1.8E-04 | | 1.00 | 0.09 |
| ABHD5 | 3 | 43732375 | 43764217 | 60 | 11 | 6.8E-06 | 4.1E-04 | 0.002 | 0.001 | 3.55 | 1.9E-04 | | 1.00 | 0.09 |

**Supplementary Table 12: Integration of hEDS GWAS with eQTL data through transcriptome-wide association (TWAS), Mendelian randomization (MR) and colocalization.**

Table indicate signficant TWAS hits (FDR<0.05) after testing association in 15 tissues based on genetic expression weight matrices computed from GTEx database (v8 release). Mendelian Randomization analysis (with eQTL as exposure and hEDS as outcome) and colocalization were performed for each identified TWAS hit. SE: Standard error. Z: Z-Score (ratio of beta over SE). IVW: inverse variance weighted. PP.H0.abf to PP.H4.abf columns respectively indicate the computed probabilities for: no association at the locus (H0), association for trait 1 (hEDS) only at the locus (H1), association for trait 2 (eQTL) only at the locus (H2), associations for both traits at the locus but with different causal variants (H3), association of both traits at the locus sharing a causal variant (H4).

| GENE | | | | | | Tissue | TWAS | | | MR | | | | | | Colocalization | | | | | |
|---|---|---|---|---|---|---|---|---|---|---|---|---|---|---|---|---|---|---|---|---|---|
| ID | Name | Chr | start | end | strand | | Z | P | FDR | Method | N SNPs | BETA | SE | P | Z | N SNPs | PP.H0.abf | PP.H1.abf | PP.H2.abf | PP.H3.abf | PP.H4.abf |
| ENSG000001 | *CFDP1* | chr16 | 75293698 | 75433503 | - | Whole_Blood | 5.00 | 5.7E-07 | 0.025 | Wald ratio | 1 | 0.92 | 0.19 | 1.5E-06 | 4.81 | 4935 | 0% | 0% | 0% | 3% | 96% |
| ENSG000001 | *CFDP1* | chr16 | 75293698 | 75433503 | - | Nerve_Tibial | 4.93 | 8.2E-07 | 0.025 | IVW | 2 | 1.09 | 0.19 | 3.7E-09 | 5.90 | 4935 | 0% | 0% | 0% | 12% | 88% |
| ENSG000001 | *MADD* | chr11 | 47269161 | 47330031 | + | Artery_Tibial | 4.82 | 1.5E-06 | 0.025 | Wald ratio | 1 | 1.42 | 0.34 | 3.7E-05 | 4.13 | 2831 | 0% | 0% | 2% | 56% | 42% |
| ENSG000001 | *CFDP1* | chr16 | 75293698 | 75433503 | - | Lung | 4.81 | 1.5E-06 | 0.025 | Wald ratio | 1 | 1.30 | 0.27 | 1.5E-06 | 4.81 | 4935 | 0% | 0% | 0% | 4% | 96% |
| ENSG000001 | *CFDP1* | chr16 | 75293698 | 75433503 | - | Heart_Left_Ventricle | 4.81 | 1.5E-06 | 0.025 | Wald ratio | 1 | 1.04 | 0.25 | 4.0E-05 | 4.11 | 4935 | 0% | 0% | 0% | 8% | 92% |
| ENSG000001 | *MADD* | chr11 | 47269161 | 47330031 | + | Whole_Blood | 4.75 | 2.1E-06 | 0.029 | Wald ratio | 1 | 2.11 | 0.47 | 6.0E-06 | 4.53 | 2831 | 0% | 0% | 0% | 13% | 87% |
| ENSG000001 | *MADD* | chr11 | 47269161 | 47330031 | + | Colon_Sigmoid | 4.62 | 3.8E-06 | 0.035 | Wald ratio | 1 | 0.40 | 0.26 | 1.2E-01 | 1.54 | 2831 | 1% | 21% | 2% | 43% | 34% |
| ENSG000001 | *CFDP1* | chr16 | 75293698 | 75433503 | - | Artery_Coronary | 4.61 | 4.1E-06 | 0.035 | Wald ratio | 1 | 1.12 | 0.23 | 1.3E-06 | 4.84 | 4935 | 0% | 0% | 0% | 8% | 92% |
| ENSG000001 | *PSMC3* | chr11 | 47418769 | 47426473 | - | Heart_Atrial_Appendage | -4.58 | 4.7E-06 | 0.035 | Wald ratio | 1 | -2.00 | 0.42 | 1.4E-06 | -4.82 | 2918 | 0% | 0% | 0% | 6% | 94% |
| ENSG000001 | *PSMC3* | chr11 | 47418769 | 47426473 | - | Skin_Not_Sun_Exposed_Suprapubic | -4.56 | 5.1E-06 | 0.035 | Wald ratio | 1 | -2.17 | 0.48 | 6.0E-06 | -4.53 | 2918 | 0% | 0% | 0% | 7% | 93% |
| ENSG000002 | *ZBED6* | chr1 | 203795714 | 203854999 | + | Skin_Sun_Exposed_Lower_leg | 4.56 | 5.1E-06 | 0.035 | Wald ratio | 1 | 1.16 | 0.27 | 1.4E-05 | 4.34 | 3921 | 0% | 0% | 1% | 9% | 90% |
| ENSG000001 | *CFDP1* | chr16 | 75293698 | 75433503 | - | Cells_Cultured_fibroblasts | 4.55 | 5.3E-06 | 0.035 | Wald ratio | 1 | 0.49 | 0.12 | 1.7E-05 | 4.30 | 4935 | 0% | 0% | 1% | 26% | 73% |
| ENSG000001 | *CFDP1* | chr16 | 75293698 | 75433503 | - | Colon_Transverse | 4.55 | 5.4E-06 | 0.035 | Wald ratio | 1 | 1.08 | 0.24 | 4.9E-06 | 4.57 | 4935 | 0% | 0% | 0% | 4% | 95% |
| ENSG000002 | *SPIRE2* | chr16 | 89818179 | 89871319 | + | Thyroid | 4.51 | 6.5E-06 | 0.037 | Wald ratio | 1 | 0.48 | 0.13 | 2.0E-04 | 3.72 | 3582 | 0% | 0% | 6% | 7% | 87% |
| ENSG000001 | *CFDP1* | chr16 | 75293698 | 75433503 | - | Heart_Atrial_Appendage | 4.51 | 6.6E-06 | 0.037 | Wald ratio | 1 | 0.96 | 0.20 | 1.5E-06 | 4.82 | 4935 | 0% | 0% | 0% | 2% | 98% |

**Supplementary Table 13: Pairwise genetic correlations between hEDS and the analysed phenotypes.**

The table reports cross-trait genetic correlations (rg) between the hEDS meta-analysis (pheno1) and external phenotypes (label_pheno2). For each phenotype we list the figure abbreviation (abbr_pheno2), clinical category (category_pheno2), the rg estimate with its standard error (se), and the two-sided P-value (pval) from bivariate LD score regression (LDSC).

Summary statistics were harmonized to the same effect allele on hg19/GRCh37, restricted to HapMap3 SNPs, and processed with ldsc's munge_sumstats.py. Analyses used European-ancestry datasets; per-trait cohort sources and sample sizes are described in Supplementary Table 7. Asterisks mark correlations that survive multiple-testing correction as specified in the Methods. Note: hernia subtypes are shown separately (inguinal, umbilical, ventral, abdominal) but are counted as one non-redundant phenotype family in multiple-testing procedures.

| pheno1 | label_pheno2 | abbr_pheno2 | category_pheno2 | rg | se | pval |
|---|---|---|---|---|---|---|
| hEDS (Meta-analysis) | Myalgic encephalomyelitis / Chronic fatigue syndrome | ME/CFS* | Pain & Autonomic | 0.35 | 0.04 | 6.47E-15 |
| hEDS (Meta-analysis) | Irritable bowel syndrome | IBS* | Gastrointestinal manifestations | 0.31 | 0.04 | 1.71E-13 |
| hEDS (Meta-analysis) | Chronic pain | CP* | Pain & Autonomic | 0.33 | 0.05 | 5.16E-12 |
| hEDS (Meta-analysis) | Migraine | MIG* | Psychiatric/Neurological | 0.30 | 0.05 | 1.95E-08 |
| hEDS (Meta-analysis) | Major depressive disorder | MDD* | Psychiatric/Neurological | 0.17 | 0.03 | 1.18E-07 |
| hEDS (Meta-analysis) | Gastroesophageal reflux disease | GERD* | Gastrointestinal manifestations | 0.18 | 0.04 | 2.87E-05 |
| hEDS (Meta-analysis) | Joint hypermobility | JH* | HEDS Spectrum | 0.41 | 0.12 | 8.00E-04 |
| hEDS (Meta-analysis) | Autism spectrum disorder | ASD | Psychiatric/Neurological | 0.13 | 0.05 | 0.008 |
| hEDS (Meta-analysis) | Gastroparesis | GAST | Gastrointestinal manifestations | 0.25 | 0.11 | 0.02 |
| hEDS (Meta-analysis) | Abdominal hernia | AH | Hernias | 0.11 | 0.05 | 0.03 |
| hEDS (Meta-analysis) | Fibromyalgia | FIB | Pain & Autonomic | 0.21 | 0.10 | 0.04 |
| hEDS (Meta-analysis) | Pelvic organ prolapse | POP | Other | 0.11 | 0.07 | 0.12 |
| hEDS (Meta-analysis) | Ventral hernia | VH | Hernias | 0.11 | 0.07 | 0.12 |
| hEDS (Meta-analysis) | Umbilical hernia | UH | Hernias | 0.07 | 0.05 | 0.19 |
| hEDS (Meta-analysis) | Anxiety disorders | ANX | Psychiatric/Neurological | 0.19 | 0.15 | 0.23 |
| hEDS (Meta-analysis) | Autonomic nervous system disorders | ANS | Pain & Autonomic | 0.09 | 0.08 | 0.24 |
| hEDS (Meta-analysis) | Tricuspid valve disease | TrVD | Circulatory | 0.14 | 0.22 | 0.51 |
| hEDS (Meta-analysis) | Mitral valve prolapse | MVP | Circulatory | 0.0211 | 0.0619 | 0.7327 |
| hEDS (Meta-analysis) | Inguinal hernia | IH | Hernias | 0.0115 | 0.0495 | 0.817 |
